# Accuracy of a Smart-Ring VO2max Estimate and Five Published Prediction Equations Against Cardiopulmonary Exercise Testing: Development and Validation Study With Population-Scale Analysis

**DOI:** 10.64898/2026.07.16.26358226

**Authors:** Nihav Dhawale, Somaaya Mukundan, Ankit Agarwal, Debasrija Mondal, Aditi Shanmugam, Pavan Kumar, Mukul Mittal, Vinayak Narasimhan

## Abstract

**Background:** Maximal oxygen uptake (VO2max) is a leading marker of cardiorespiratory fitness and a strong predictor of all-cause mortality. Cardiopulmonary exercise testing (CPET) is the reference method but is resource-intensive, so consumer wearables estimate VO2max from passively collected signals; these estimates compress the fitness range, returning near-correct group averages while ranking individuals poorly. No peer-reviewed validation of a smart-ring VO2max estimate against CPET has been reported, and none in a South Asian cohort.

**Objective:** To validate the Ultrahuman Ring AIR VO2max estimate against laboratory CPET, benchmark it against published prediction equations, and assess its generalization and construct validity.

**Methods:** In a single-site paired ring-CPET cohort (N=101; CPET peak VO2 43.3 mL·kg⁻¹·min⁻¹, SD 9.9), peak oxygen uptake was measured by treadmill or cycle-ergometer CPET, and the Ultrahuman Ring AIR estimate was computed from passively collected signals using a transparent ensemble based on published equations. Ensemble weights and calibration were selected on an 85-subject development set by an automated search minimizing a composite 5-fold cross-validated error criterion; the locked estimate was evaluated on a 16-subject held-out test set. The calibrated coefficients are proprietary. Agreement was quantified with mean absolute error (MAE), bias, Pearson r, regression slope and Lin’s concordance correlation coefficient (CCC; bootstrap 95% CIs), and Bland-Altman limits of agreement. Separately, in 181,133 deidentified Ring users (no CPET reference), construct validity was assessed against ring-measured sleep, continuous glucose monitoring (n=2,597), and a venous blood panel (n up to 15,203), adjusted for age, sex, and BMI, with lipoprotein(a) as a pre-specified negative control. Reporting followed TRIPOD and STARD.

**Results:** With a self-reported fitness level provided, the estimate agreed with CPET peak VO2 at MAE 4.68 mL·kg⁻¹·min⁻¹ (95% CI 3.93 to 5.49), Pearson r 0.79, CCC 0.79, and slope 0.71. The five published equations were worse on every metric (MAE 6.2 to 10.6, CCC 0.28 to 0.56, slope 0.32 to 0.42), each compressing the fitness range. On the held-out test set (n=16), agreement held (r 0.84, slope 0.81, MAE essentially unchanged). Without the fitness input, full-cohort MAE was 5.16, still ahead of every published equation. At population scale, higher estimated fitness tracked a healthier profile on measurements the estimate does not use: better ring-measured sleep; higher continuous-glucose time in target range (79.6% versus 61.5%, top versus bottom decile; n=222 and 399 of 2,597 users); and lower triglycerides, fasting glucose, and HOMA-IR (n up to 15,203 assayed per marker). These associations held after adjustment for age, sex, and BMI, whereas the pre-specified negative control lipoprotein(a) did not separate the deciles.

**Conclusions:** The Ultrahuman Ring AIR VO2max estimate agreed with laboratory CPET substantially better than published prediction equations, held its agreement on held-out subjects, and ordered a large population along independent cardiometabolic gradients consistent with true fitness.

**Trial Registration:** Not applicable.

## Introduction

Maximal oxygen uptake (VO2max) is the integrated measure of the cardiorespiratory system’s capacity to deliver and use oxygen during exercise, and it is among the strongest markers of long-term health. In a meta-analysis of healthy adults, each 1-MET increment in fitness (one metabolic equivalent of task) was associated with roughly 13% lower all-cause mortality [1], and in more than 120,000 adults referred for treadmill testing the lowest-fitness group carried about five times the mortality of the elite-fitness group [2]. On this evidence the American Heart Association has argued that cardiorespiratory fitness be treated as a clinical vital sign [3]. The reference method is cardiopulmonary exercise testing (CPET), a graded test to volitional exhaustion with breath-by-breath gas exchange; it is accurate but resource-intensive, requiring a metabolic cart, a supervised maximal effort, and trained staff, so it is rarely available outside the laboratory [3,4]. That access gap has motivated consumer wearables to estimate VO2max from passively collected signals.

Wearable VO2max estimates share a recurring weakness: they track the population mean acceptably while compressing the individual range, so low-fitness users are over-read and high-fitness users under-read. The INTERLIVE systematic review and expert consensus concluded that accuracy is reasonable at the group level while individual-level error remains substantial [5]. The signature of this error is a regression slope below one against the reference, often with a near-zero mean bias that hides the compression when only group means are inspected; it recurs across manufacturers and form factors, including single-site evaluations of flagship smartwatches [6]. When an estimate orders individuals poorly, it undermines the marker’s central purpose, the one its mortality association implies: identifying who is at low fitness.

These evaluations have centered on wrist- and chest-worn devices. Smart rings have become a distinct class of wearable, but no ring VO2max output has been evaluated against CPET. Peer-reviewed validation of smart-ring outputs has reached sleep duration and resting heart rate [7], sleep staging [8], and heart-rate variability [9], but no ring VO2max estimate has been validated against a reference test [7]. Furthermore, reviews that catalogue wearable VO2max accuracy by sensing modality include wrist, chest, foot-pod, and power-meter systems but not a ring [10]. The cohorts studied are a second gap: prior wearable VO2max validations were conducted in European, North American, and East Asian populations [5,6], with none in a South Asian cohort, a population in which laboratory measurement has documented lower mean fitness at an equivalent body-mass index than in European men [11]. Prior work in Indian cohorts has estimated VO2max from heart-rate-based prediction models validated against laboratory exercise testing [12], but did not evaluate a consumer ring’s native output against CPET.

To our knowledge, this study reports the first validation of a smart-ring VO2max estimate, the Ultrahuman Ring AIR, against laboratory CPET, and the first in an Indian cohort, a South Asian population underrepresented in prior wearable validations. The aims are threefold: to quantify agreement using metrics that expose range compression (regression slope and Lin’s CCC) rather than correlation and group-mean error alone; to benchmark that agreement against five published non-exercise and heart-rate prediction equations on the same cohort; and to assess whether the estimate generalizes to held-out subjects and how it behaves at population scale, including its construct validity against independent cardiometabolic measures.

## Methods

### Study design and participants

This was a single-site, retrospective observational analysis pairing a smart-ring VO2max estimate with laboratory CPET as the reference standard. It used de-identified records from adults who underwent a paired CPET-and-ring assessment at the Ultrahuman Performance Lab (Bangalore, India), each contributing one CPET measurement and concurrent ring wear. Eligibility required an age of 18 years or older, a valid CPET peak VO2 measurement, and a concurrent ring-derived night-time resting heart rate; the latter two are the inputs the agreement analysis depends on.

Demographics (age, biological sex, body weight, height) were recorded at the visit, and BMI was computed as weight in kilograms divided by height in metres squared. A mobility (activity) tier was derived from the 7-day average daily step count, and a self-reported fitness level was recorded when supplied. The analysis cohort comprised 101 evaluable adult participants spanning a wide fitness range, which supports agreement analysis across the fitness distribution rather than within a narrow band (full demographics in Results, Table 1). CPET assessments were conducted between January 11 and June 10, 2026. Participants attended the laboratory in two series: 24 in an earlier series tested between January 11 and February 27, 2026, and 77 recruited through an open-call VO2max Challenge campaign for the laboratory’s assessment service and tested between June 1 and 10, 2026. The cohort is therefore a self-selected convenience series rather than a consecutive one.

**Table 1.** Participant characteristics (n=101)

| Variable | Value |
| --- | --- |
| Participants, n | 101 |
| Sex | 72 male (71.3%), 29 female (28.7%) |
| Age, y | 32.6 (7.5), 19-56 |
| Weight, kg | 68.9 (12.8), 44.9-110.2 (n=101) |
| Height, cm | 170.2 (8.8), 150-186 (n=99)<sup>b</sup> |
| BMI, kg·m<sup>-2</sup> | 23.7 (3.1), 16.9-33.8 (n=99)<sup>b</sup> |
| Mobility tier<sup>a</sup> | 38 low (37.6%), 42 medium (41.6%), 21 high (20.8%) |
| Self-reported fitness | 34 Advanced, 18 Intermediate, 7 Elite, 4 Beginner (n=63)<sup>c</sup> |
| CPET peak VO<sub>2</sub>, mL·kg<sup>-1</sup>·min<sup>-1</sup> | 43.3 (9.9), 21.6-66.1 |
| CPET peak VO<sub>2</sub> by sex, mL·kg<sup>-1</sup>·min<sup>-1</sup> | male 45.4 (10.3), n=72; female 38.0 (6.0), n=29 |

Values are mean (SD), range, unless otherwise indicated. CPET indicates cardiopulmonary exercise test; BMI, body mass index; peak VO<sub>2</sub>, peak oxygen uptake (VO<sub>2</sub>peak).
<sup>a</sup> Mobility tier is the device-derived activity-mobility stratum used by the algorithm.
<sup>b</sup> Height was missing for 2 participants: one was never recorded, and one recorded value (127 cm) fell outside the adult plausibility bound of 140 to 210 cm and was set to missing as a data-entry error (Methods; Multimedia Appendix 1, §1.3). BMI is therefore computed on n=99.
<sup>c</sup> Self-reported fitness level was provided by 63 of 101 participants (38 missing); this optional input conditions the Ultrahuman estimate and is the reason the default deployed path scores all users at the neutral central fitness setting (the "everyone-medium" default, which is distinct from the low/medium/high mobility tier above; see Table 3 and Limitations).

To keep a data-entry error from entering the anthropometric inputs, height was screened against a fixed adult plausibility bound of 140 to 210 cm. The bound was set from the biology rather than from the data and was applied to every record without reference to the CPET result or to any estimator’s error. One recorded height, 127 cm at a recorded weight of 69 kg, fell outside it. A height of 127 cm is not a possible adult measurement, and the bound sits 10 cm below the shortest genuine height in the cohort (150.0 cm), so no real measurement is at risk of being caught by it. That height, and the BMI derived from it, were set to missing; the participant was retained, with every other input unchanged. One further participant had no height on record, so height was missing for 2 of the 101 participants. Each estimator then applied its own rule for a missing height (Multimedia Appendix 1, §1.2). A sensitivity analysis that instead retains the erroneous value, and in which no conclusion changes, is reported in Multimedia Appendix 1, §1.3.

### Ethical Considerations

The study was conducted in accordance with the Declaration of Helsinki (2013 revision). The analysis of the de-identified paired CPET-and-ring dataset, and its planned publication, were reviewed and approved by the Medstar Speciality Hospital Ethics Committee, Bangalore, India (CDSCO Ethics Committee Registration No. ECR/1324/Inst/KA/2019/RR-24), by letter dated 04 July 2026. Both the paired ring-CPET analysis and the population-scale analysis of de-identified Ultrahuman platform records were approved under this review as retrospective observational research under the Indian Council of Medical Research (ICMR) National Ethical Guidelines (2017); neither analysis is a clinical trial under the New Drugs and Clinical Trials Rules (2019), so no prospective trial registration applies. Participants in the paired ring-CPET cohort had given written informed consent for laboratory testing, concurrent wearable data collection, and use of their de-identified data for research, under the Ultrahuman Performance Lab master consent form and, for those recruited through the VO2max Challenge campaign, an additional VO2max Challenge consent form. Participants whose images appear in this manuscript gave written consent for the publication of those images. Participants were not recruited for research and received no compensation for research participation; they attended the Performance Lab for a fitness assessment, and the analysis reported here is retrospective on their de-identified records. The population-scale analysis used de-identified Ultrahuman platform records across three channels, the ring, the continuous glucose monitor, and the venous blood panel, all collected under the Ultrahuman Privacy Policy and Terms of Service, which permit the analysis of de-identified grouped data for scientific research and to which users consent at onboarding and through continued use of the platform. For these retrospective components the Ethics Committee approved a waiver of additional consent under the ICMR National Ethical Guidelines (2017, §5.5); all records were de-identified before analysis.

### Reference test: cardiopulmonary exercise testing

CPET served as the reference standard, providing peak oxygen uptake as the reference value. Each participant performed a graded treadmill or cycle-ergometer test to volitional exhaustion with breath-by-breath gas exchange measured by a metabolic cart, and peak oxygen uptake (VO2peak, mL·kg⁻¹·min⁻¹), defined as the highest 30-second rolling average of oxygen uptake, was taken as the reference value. Of the 101 participants, 78 were tested on the treadmill and 21 on the cycle ergometer; the source test record did not carry the modality for the remaining 2. The ring was worn during the visit and surrounding days so the ring-derived estimate corresponded to the same measurement period as the CPET assessment. Both treadmill and cycle-ergometer tests used the same stationary metabolic cart (Quark CPET; COSMED, Rome, Italy), running OMNIA software (version 2.4.2). The gas analysers were calibrated each day before the first test against a certified reference gas mixture, flow and volume were calibrated each week with a 3-L syringe, and ambient temperature, humidity, and barometric pressure were recorded before each test and applied in the gas-exchange calculations. Treadmill testing followed a ramp protocol: a 3-minute warm-up at 3 km/h and 0% grade, then a constant 1% grade with speed starting at 4 km/h and increasing 0.5 km/h every 30 seconds to volitional exhaustion, followed by a 5-minute cool-down at 3 km/h. Cycle-ergometer testing used an individualized ramp whose rate was derived from the prediction equation of Jones and Makrides [13] and rounded to the nearest 5 W/min, with a 3-minute warm-up at 20 W (45 to 55 rpm), a continuous ramp from 20 W to the limit of tolerance, and a 5-minute cool-down at 25 W. Participants were verbally encouraged to volitional exhaustion, and effort was assessed against standard maximal-effort indicators: a respiratory exchange ratio approaching or exceeding 1.10, a peak heart rate near the age-predicted maximum, and a plateau in oxygen uptake where present. As is common in cardiopulmonary exercise testing, not every participant reached a respiratory exchange ratio of 1.10 or showed an oxygen-uptake plateau, so the highest attained oxygen uptake (VO2peak) was retained as the reference for every participant rather than a plateau-confirmed VO2max. No adverse events occurred during any test.

### The Ultrahuman VO2max estimate

The Ultrahuman Ring AIR VO2max estimate reports VO2max (mL·kg⁻¹·min⁻¹) from passively collected signals, without a maximal or supervised exercise test. We follow the disclosure convention of comparable consumer metrics, Apple’s Cardio Fitness white paper [14] and the Firstbeat/Garmin VO2max white papers [15,16]: we specify the method’s inputs, structure, and calibration philosophy, and hold the final calibrated coefficients and ensemble weights proprietary. This level of disclosure lets the approach be understood and critiqued without being reproduced commercially.

The estimate uses night-time resting heart rate (RHR) from photoplethysmography during sleep, demographics (age, sex, weight, height, combined as BMI where used), daily ambulatory activity (step count and a derived mobility tier), and an optional self-reported fitness level that refines the estimate when present and otherwise falls back to a population default. The point estimate is not a single black-box regressor but a weighted sum of three terms, each following the functional form of an independently published prediction equation and capturing a complementary pathway: a heart-rate-ratio term following the Uth relationship [17], a heart-rate-reserve term in the ACSM submaximal tradition [18], and a non-exercise demographic regression in the Jackson tradition on age, sex, BMI, and activity [19]. Each component is identified with its published source in Multimedia Appendix 1 (§1.1); the coefficients within each component were re-fitted on the development set. Maximum heart rate comes from an age-prediction equation [20] deliberately decoupled from same-day activity to avoid a mobility-trap bias in which low daily movement depresses apparent achievable heart rate and inflates the estimate. The ensemble output is adjusted by a population-level calibration offset, a policy-tunable population dial rather than a per-user fit that aligns the cohort distribution to reference-laboratory norms, and stabilized across days with exponential-moving-average (EMA) smoothing (Figure 1). We disclose the input set, the identity and physiological meaning of each component, the max-HR decoupling, the calibration offset, and the EMA smoothing; we declare proprietary the ensemble weights, the calibrated coefficients and offset magnitude, the activity-tier multipliers, and the smoothing constant. The ensemble weights and the component coefficients were not set by hand: they were selected by an automated configuration search on a development subset of the cohort, described in the Statistical analysis and in Multimedia Appendix 1 (§1.1), where the max-HR decoupling is also specified. Every estimate reported here was recomputed from each participant’s stored ring signals with a single locked build of the algorithm (frozen June 23, 2026), so one identical configuration scored the whole cohort.

**Figure 1.**
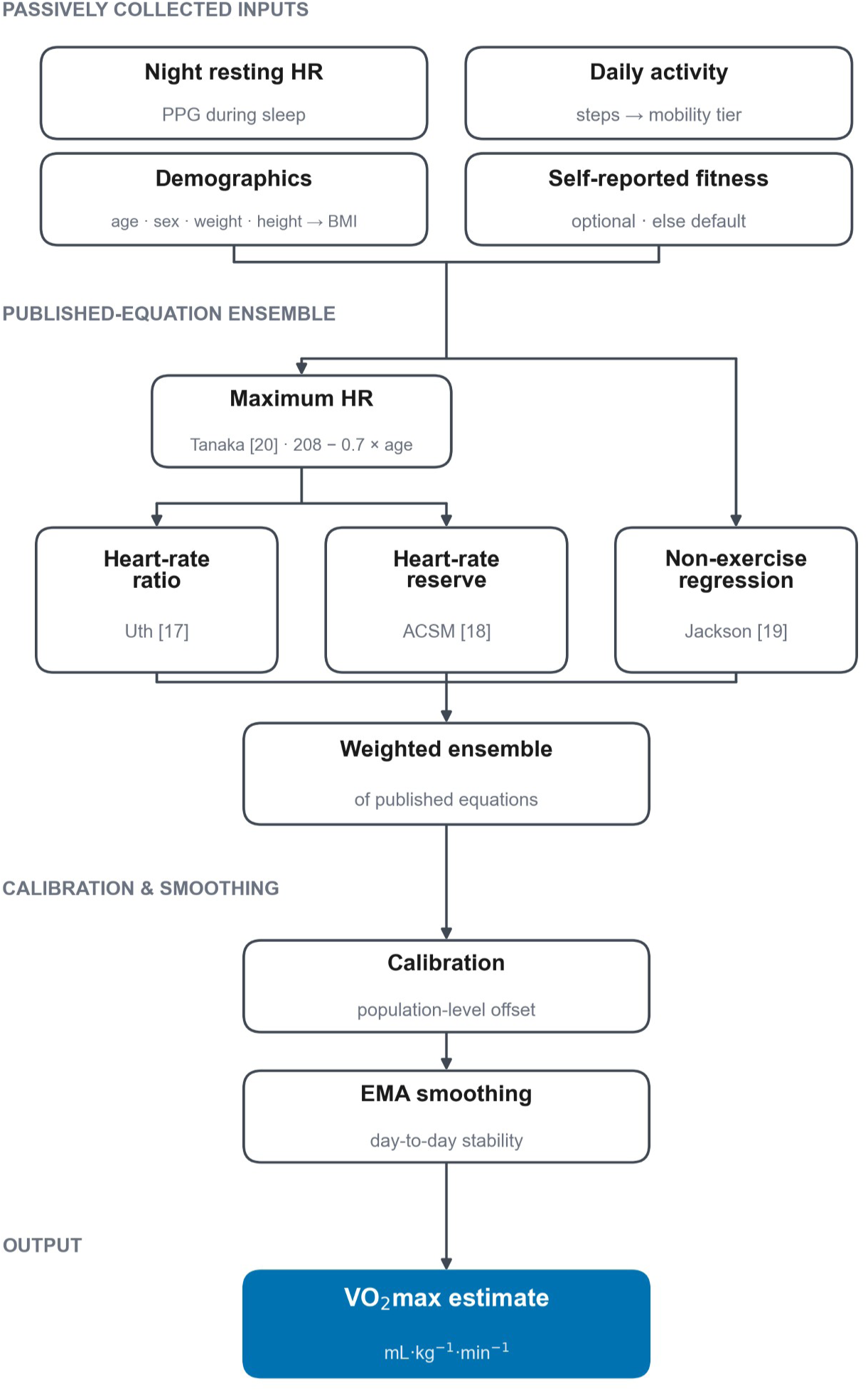
Schematic of the Ultrahuman VO2max estimation algorithm. Passively collected inputs (night-time resting heart rate from photoplethysmography during sleep; demographics, combined as BMI; daily activity as a step-derived mobility tier; and an optional self-reported fitness level that otherwise defaults to a population value) feed a transparent weighted ensemble of three terms, each following the functional form of a published prediction approach: a heart-rate-ratio term (Uth [17]), a heart-rate-reserve term (ACSM submaximal tradition [18]), and a non-exercise demographic regression (Jackson [19]). Maximum heart rate is taken from the Tanaka age equation [20] and decoupled from same-day activity. The ensemble output is then adjusted by a population-level calibration offset and stabilized across days by exponential-moving-average smoothing to yield the VO2max estimate. The schematic shows the structure only; the ensemble weights, calibrated coefficients, calibration-offset magnitude, activity-tier multipliers, and smoothing constant are proprietary (Methods).

### Comparator estimators

To place the estimate against the published literature, we computed five established non-exercise and heart-rate prediction equations on the same cohort: Uth [17], Jurca [21], Jackson [19], the FRIEND registry reference equation (de Souza e Silva) [22], and HUNT/Nes [23]. Each received the same ring-derived inputs as the Ultrahuman estimate. Because a wearable infers some quantities the original equations measured directly or by questionnaire (resting heart rate, an activity questionnaire, waist circumference), each equation was applied with disclosed input approximations carried uniformly across methods, including the ring’s night-time sleeping RHR (a few beats·min⁻¹ below clinic RHR) wherever a resting heart rate was required and a tier-specific age-prediction equation for maximum heart rate where one was needed [24,25,26]; the per-equation specifics are in Multimedia Appendix 1 (§1.2).

### Statistical analysis

CPET peak VO2 was determined by the Performance Lab independently of the ring data. The estimate for each participant is computed from that participant’s ring signals alone, without access to their CPET value; the algorithm’s ensemble weights and calibration, however, were selected on the development set’s CPET values (described below), so agreement on the full cohort partly reflects in-sample fit, and the held-out test set provides the out-of-sample estimate. No a-priori sample-size calculation was performed; all available paired assessments were included, and achieved precision is reported as bootstrap confidence intervals around each metric.

The primary analysis quantified agreement on the full cohort (n=101). To capture both error magnitude and the structure of disagreement we report mean absolute error (MAE) and mean bias (estimate minus reference); Pearson r and the slope of the ordinary-least-squares (OLS) regression of estimate on CPET, where a slope of 1 indicates no range compression and a slope below 1 indicates compression; Lin’s concordance correlation coefficient (CCC), which, unlike r, is reduced by systematic under- or over-estimation [27]; and the proportion of estimates within 5 mL·kg⁻¹·min⁻¹ of the reference. Agreement is also summarized by the Bland-Altman method, with mean difference and 95% limits of agreement (bias ± 1.96 SD) [28]. We designate the regression slope and Lin’s CCC as the headline agreement metrics, treating the other measures as supporting, because a near-zero bias and a moderate correlation can coexist with range compression that only slope and CCC expose. Bootstrap 95% confidence intervals used 5000 resamples of participants with replacement (fixed seed 42).

The estimate’s ensemble weights and calibration were developed on an 85-subject development set, defined as a random split of the cohort; the roughly 85%/15% development/held-out proportion is a conventional choice for hold-out validation, and the repeated, seed-averaged cross-validation within the development set was used precisely so the configuration would not hinge on any single fold assignment. Development used an automated configuration search (an “autoresearch” loop, described in the Use of Artificial Intelligence section): the search repeatedly proposed a candidate configuration of ensemble weights, calibration, and component coefficients, scored it, and kept the change only when it lowered a composite error criterion dominated by repeated 5-fold cross-validated mean absolute error on the development set (the full procedure, the search objective, and the development/held-out split are detailed in Multimedia Appendix 1, §1.1). The locked configuration was then evaluated on a held-out test set of 16 subjects that was excluded from the cross-validation folds and from the search keep/discard loop; this set was consulted once to choose between two final candidate configurations and then used to evaluate the locked estimate, so it provides a near-out-of-sample (quasi-independent) read rather than a strictly untouched test set (Multimedia Appendix 1, §1.1). For the held-out set we lead with correlation and slope, which test whether the estimate’s ordering and range agreement transfer to unseen participants, and we state its n each time given its small size. Because the headline accuracy uses the optional self-reported fitness input, we also computed agreement under the default deployed configuration, in which that input is absent and every participant is scored on the medium tier, across the same splits.

Next, a population-scale analysis characterized deployment behavior on the default configuration: each method was applied to a 181,133-user cohort, computing one per-user estimate from per-user mean inputs. Because there is no CPET reference at population scale, this analysis is distributional only (median, interquartile range, mean, and the proportion of estimates at the low-fitness floor of ≤16 mL·kg⁻¹·min⁻¹); no agreement metric or slope is reported there. We report this study against TRIPOD as the primary reporting guideline, because the estimate is a multivariable prediction model [29]. For the Ultrahuman estimate the study is a development and validation using a random split-sample of a single cohort (TRIPOD type 2a), not an external validation: the ensemble weights and calibration were selected on the 85-subject development set, and the locked configuration was then evaluated on the 16 held-out subjects. For the five published equations, which are applied as published with no fitting to this cohort, the study is a validation of existing models (TRIPOD type 4). No external validation of the Ultrahuman estimate in an independent population is presented. We report against STARD as a secondary guideline because CPET is the reference standard [30]; completed checklists are provided as Multimedia Appendix 2 (TRIPOD) and Multimedia Appendix 3 (STARD).

Analysis used plain numerical computation in Python 3.12.12 (NumPy 2.4.3, SciPy 1.17.1, pandas 3.0.1, statsmodels 0.14.6); the deployed estimate is a closed-form ensemble of published equations and contains no learned black-box model. Analysis and figure scripts are available on reasonable request (Data Availability).

### Population correlate analysis

To assess construct validity beyond the paired corpus, we examined how the validated estimate orders a large population against measurements it does not use. We reviewed 181,133 Ultrahuman Ring AIR de-identified users with at least 20 nights of valid night-time RHR in a fixed recent 90-day window and complete age, sex, height, and weight (124,886 female, 56,247 male; mean age 34.8 years), and recomputed the estimate for each user on the default medium-tier path. Because the estimate depends on sex and age, we removed both before ranking: within each sex we regressed the estimate on age and ranked users by the residual, that is, by how far each user’s estimate sat above or below the value expected for their age and sex. The bottom and top deciles of that residual defined the low- and high-fitness groups, which are balanced on age and sex by construction. We then compared these two groups on measurements drawn from the ring itself, a continuous glucose monitor, and a venous blood panel, most of which are not inputs to the estimate. The correlates were grouped by how independent each is from the estimate: the algorithm’s own inputs (positive controls); ring sleep architecture the algorithm does not use; and two channels measured by separate instruments, continuous glucose monitoring (Ultrahuman M1 CGM; n=2,597) and venous blood markers (Ultrahuman Blood Vision; up to n=15,203 per marker). Lipoprotein(a) was pre-specified as a negative control because it is genetically determined and not lowered by habitual exercise [31]. Each correlate was compared between deciles with the Mann-Whitney test (median difference with a 1000-sample bootstrap 95% CI and the rank-biserial effect size), and its association with the continuous estimate was re-estimated by OLS regression of the standardized correlate on the standardized estimate adjusted for age, sex, and BMI; the standardized between-decile difference (Cohen’s d) is the headline effect size. Adjusting for BMI is conservative because BMI is itself an input to the estimate, so the adjusted coefficient isolates the association not carried by the body-mass term. P values were corrected within each correlate group by the Benjamini-Hochberg procedure (FDR). Decile membership was stable across estimators (Spearman ρ = 0.80). This analysis is cross-sectional. Full procedures, the estimator-stability check, and per-layer results are in Multimedia Appendix 1 (§3).

## Results

### Cohort and peak VO2 distribution

The paired ring-CPET corpus defines the population in which every agreement metric below is computed. The 101 participants were 72 male (71.3%) and 29 female (28.7%), with a mean age of 32.6 years (SD 7.5) and a mean BMI of 23.7 kg·m⁻² (SD 3.1; n=99). Ring-derived mobility tier was low in 38 (37.6%), medium in 42 (41.6%), and high in 21 (20.8%). A self-reported fitness level was present for only 63 of 101 participants; the remaining 38 were scored on the medium tier, the default the deployed estimate applies when this optional input is absent. CPET peak VO2 averaged 43.3 mL·kg⁻¹·min⁻¹ (SD 9.9) and spanned 21.6 to 66.1, roughly a threefold range from low to high fitness, with the expected sex difference (Table 1). That wide spread is what makes slope and CCC informative, since both are tested across the full fitness range. The cohort composition and CPET peak VO2 distribution are shown in Figure 2b, 2c; the estimation algorithm these are evaluated against is summarized in Figure 1.

**Figure 2.**
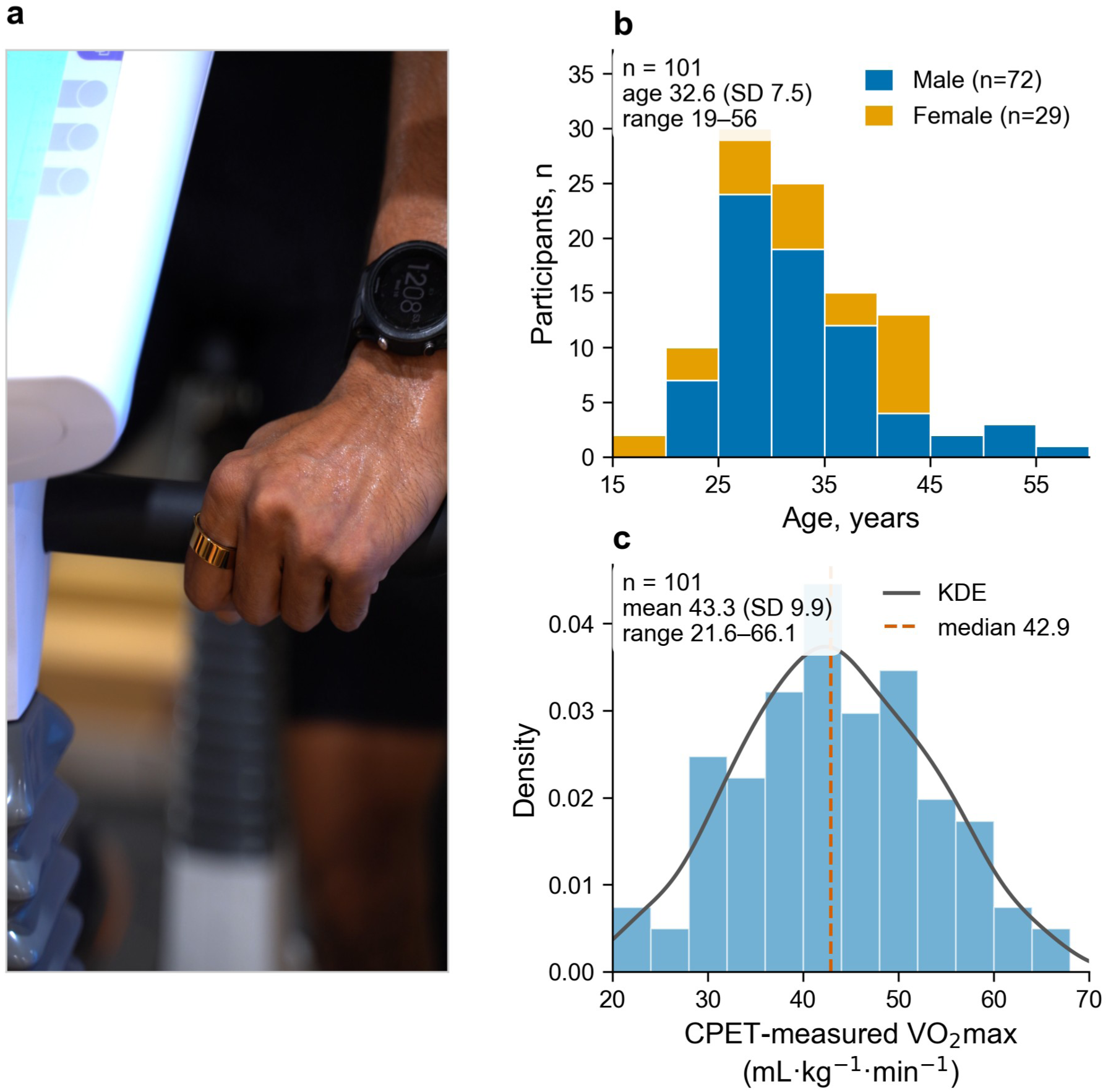
Data collection, cohort composition, and reference measurement. (a) A participant wearing the Ultrahuman Ring AIR during the cardiopulmonary exercise test. (b) Cohort composition: age distribution by sex (72 male, 29 female; mean age 32.6 years, SD 7.5). (c) Distribution of CPET-measured VO2max (histogram with kernel-density overlay and median line; n=101, mean 43.3 mL·kg⁻¹·min⁻¹, SD 9.9, range 21.6-66.1).

### Agreement with CPET

The primary analysis quantified how closely each estimator reproduced laboratory CPET across the full cohort (n=101), with a self-reported fitness level provided. This cohort includes the development subjects on which the estimate was tuned, so it is the full-sample fit; the out-of-sample read follows in the next section. The Ultrahuman Ring AIR estimate agreed with CPET-measured peak VO2 at MAE 4.68 mL·kg⁻¹·min⁻¹ (95% CI 3.93 to 5.49), Pearson r 0.792 (95% CI 0.715 to 0.855), slope 0.708 (95% CI 0.581 to 0.838), and 60.4% of estimates within 5 mL·kg⁻¹·min⁻¹; Lin’s CCC was 0.787 (95% CI 0.703 to 0.847) [27], and the Bland-Altman bias was −0.40 mL·kg⁻¹·min⁻¹ (95% CI −1.61 to +0.80) with 95% limits of agreement [−12.36, +11.56] [28]. The five published equations clustered at MAE 6.2 to 10.6, r 0.61 to 0.69, CCC 0.28 to 0.56, and slope 0.32 to 0.42, with mean biases from −2.30 to +9.90 and correspondingly wider limits of agreement (Table 2; Figure 3 and Figure 4a, 4b; per-formula scatters in Figure S1).

**Figure 3.**
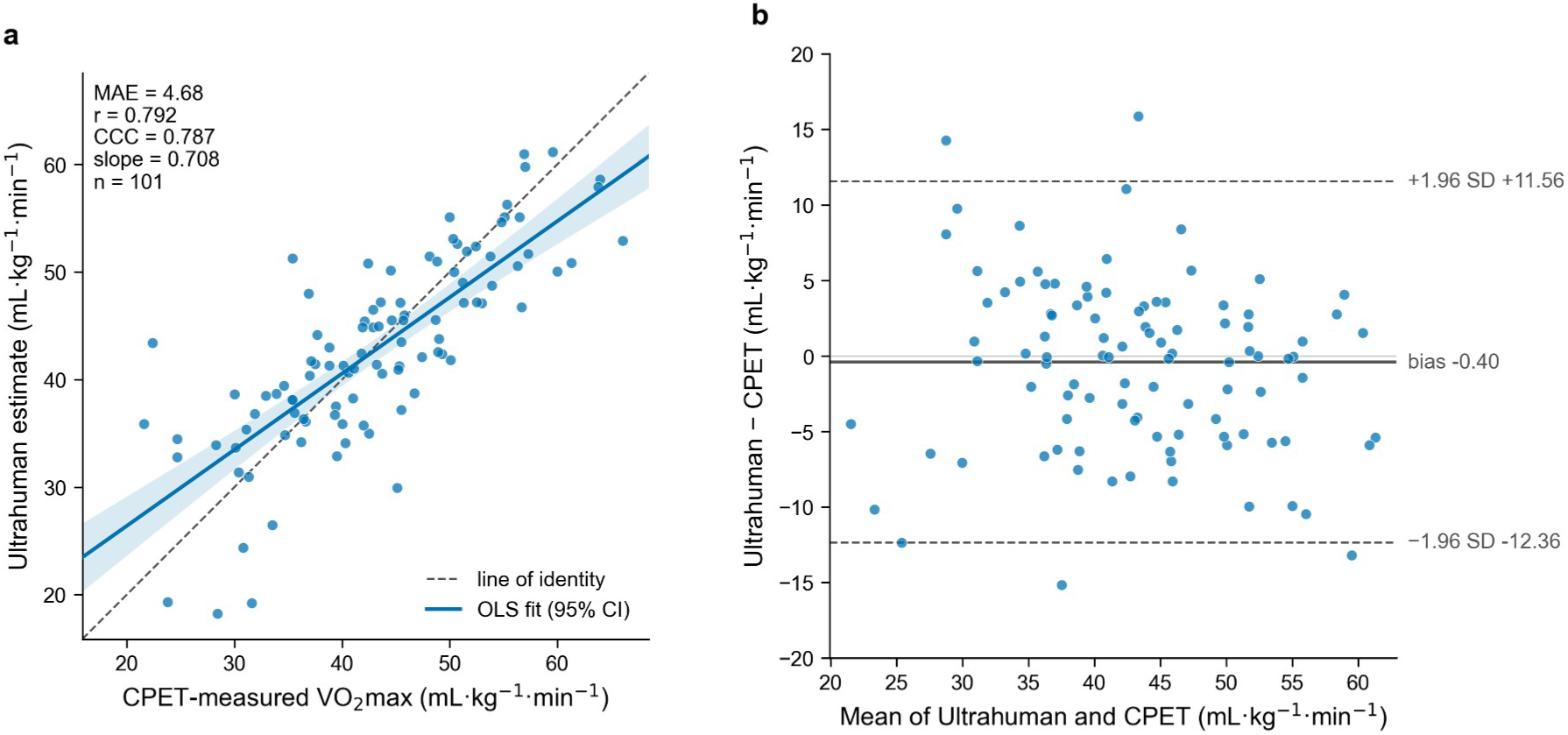
Agreement between the Ultrahuman Ring AIR estimate and CPET-measured VO2max, with a self-reported fitness level provided (n=101). (a) Agreement scatter of the Ultrahuman estimate against CPET-measured VO2max with the line of identity (dashed) and the ordinary-least-squares fit with 95% confidence band (r=0.792, Lin CCC=0.787, MAE=4.68, slope=0.708). (b) Per-subject Bland-Altman plot of the difference (estimate minus CPET) against the mean, with the mean bias (−0.40) and 95% limits of agreement (−12.36, +11.56).

**Figure 4.**
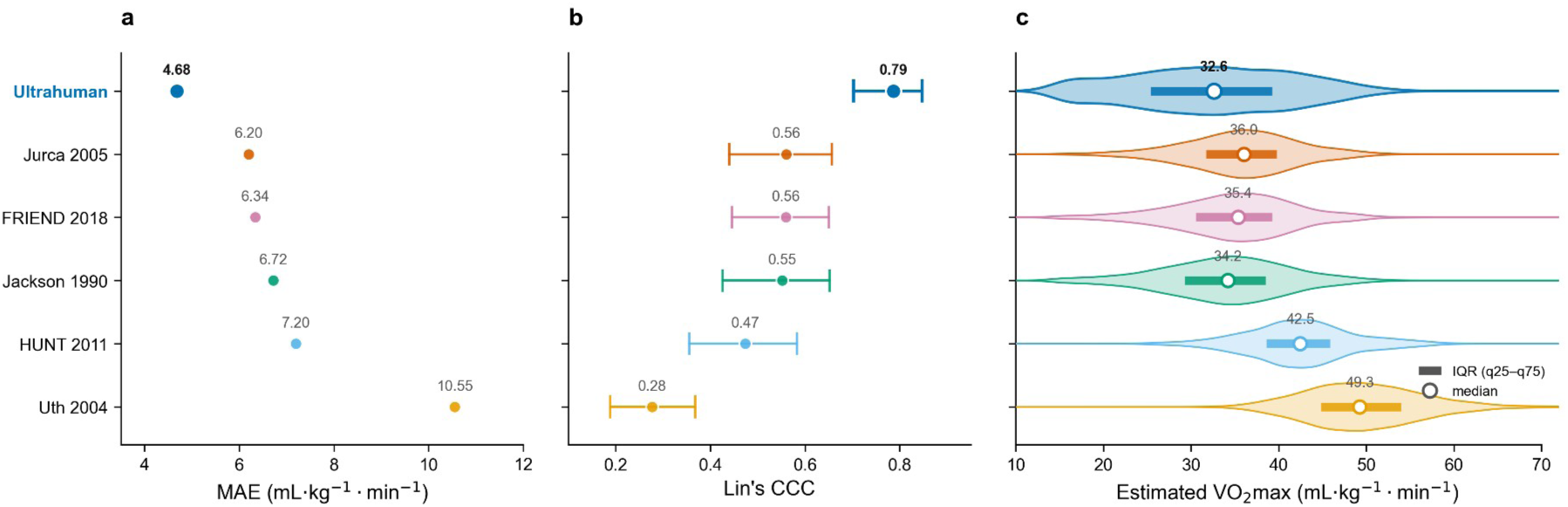
Comparison with published prediction equations: benchmark of the Ultrahuman Ring AIR estimate against five published non-exercise and heart-rate prediction equations (Uth, Jurca, Jackson, FRIEND, and HUNT/Nes), across six methods in all. Panels (a) and (b) are CPET agreement against the measured reference standard (n=101, ground truth); panel (c) is the population landing distribution, for which there is no reference standard at scale. (a) Mean absolute error and (b) Lin concordance correlation coefficient for the Ultrahuman estimate and the five published equations, with CCC whiskers as bootstrap 95% confidence intervals (5000 resamples). (c) Population-scale landing distribution, one horizontal violin per method with the median (open circle) and interquartile range (bar), on the default everyone-medium configuration; distributional only, with no CPET ground truth at population scale (n=181,133 users, the same cohort as the construct-validity analysis). Methods are ordered by ascending MAE in every panel and the Ultrahuman row is highlighted.

**Table 2.** Per-method agreement with cardiopulmonary exercise testing (n=101)

| Method | MAE | Pearson r | OLS slope (95% CI) | Within 5 mL·kg<sup>-1</sup>·min<sup>-1</sup>, % | Lin CCC (95% CI) | Bland-Altman bias | 95% LoA |
| --- | --- | --- | --- | --- | --- | --- | --- |
| Ultrahuman<sup>a</sup> | 4.68 | 0.792 | 0.708 (0.581 to 0.838) | 60.4 | 0.787 (0.703 to 0.847) | -0.40 | [-12.36, +11.56] |
| Jurca 2005 | 6.20 | 0.645 | 0.381 (0.289 to 0.474) | 45.5 | 0.560 (0.440 to 0.656) | -1.05 | [-15.88, +13.77] |
| FRIEND 2018 | 6.34 | 0.631 | 0.392 (0.304 to 0.480) | 42.6 | 0.560 (0.446 to 0.650) | -1.20 | [-16.21, +13.81] |
| Jackson 1990 | 6.72 | 0.615 | 0.419 (0.311 to 0.526) | 37.6 | 0.552 (0.426 to 0.653) | -2.30 | [-17.61, +13.01] |
| HUNT/Nes 2011 | 7.20 | 0.687 | 0.420 (0.333 to 0.509) | 48.5 | 0.474 (0.356 to 0.583) | +6.20 | [-7.94, +20.33] |
| Uth 2004 | 10.55 | 0.612 | 0.317 (0.230 to 0.411) | 23.8 | 0.277 (0.189 to 0.368) | +9.90 | [-5.51, +25.31] |

CPET peak VO<sub>2</sub> is the reference standard. MAE, Bland-Altman bias, and 95% limits of agreement (LoA) are in mL·kg<sup>-1</sup>·min<sup>-1</sup>. Bland-Altman bias is the mean signed difference (estimate minus CPET). CCC indicates Lin concordance correlation coefficient. 95% CIs for the OLS slope and Lin CCC were obtained by bootstrap resampling of participants (5000 resamples, seed 42); the limits of agreement are the mean difference ± 1.96 SD of the differences.
<sup>a</sup> The Ultrahuman estimate is reported with a self-reported fitness level provided (conditioned path); the self-reported level was used for the 63 participants who provided one, and the remaining 38 were scored on the medium default in this path as well, so the conditioned-versus-everyone-medium difference reflects those 63. The default deployed path, which does not require this optional input, is reported in Table 3 (everyone-medium default).

The contrast turns on slope and CCC rather than bias and group-mean error. Each published formula reads high or compresses the range: the Uth heart-rate-ratio method overestimated most (bias +9.90, CCC 0.277) [17], while the Jurca [21], Jackson [19], FRIEND [22], and HUNT/Nes [23] models clustered at MAE 6.2 to 7.2, none exceeding a CCC of 0.560 despite Pearson r in the 0.61 to 0.69 range. The Ultrahuman estimate raised the slope of the best published equation by two-thirds (0.42 to 0.708) and lifted CCC to 0.787, so its advantage is on the concordance axis, not merely on mean error.

### Out-of-sample generalization

Because the estimate’s weights and calibration were selected on the development set, the full-cohort agreement above partly reflects in-sample fit; a cleaner read of performance comes from data outside the development set. Generalization was tested two ways, both on random splits: 5-fold cross-validation within the 85-subject development set, and a held-out test set of 16 subjects excluded from the cross-validation folds and from the search. Because the repeated cross-validated MAE was itself the search objective, the cross-validation figures reflect in-development optimization rather than independent generalization; the held-out set, consulted only once to choose between two final configurations, is the nearest to an out-of-development read. Cross-validation reached r 0.776 and slope 0.682 (95% CI 0.544 to 0.822; MAE 4.61, 59.9% within 5), and on the held-out set r was 0.836 and slope 0.807 (95% CI 0.455 to 1.170; MAE 5.54, 56.3% within 5), so range agreement held on subjects outside the development set, while MAE stayed essentially flat; the two slope intervals overlap, so the lift is a consistent direction rather than an established difference (Table 3; Figure 5). The development (n=85) and held-out (n=16) subsets were balanced on the outcome: CPET peak VO2 averaged 43.1 mL·kg⁻¹·min⁻¹ (SD 9.5) in the development subset and 44.3 (SD 11.8) in the held-out subset (two-sample t test, P=.65), with matched fitness-band composition (low 20.0% vs 18.8%, medium 36.5% vs 37.5%, high 43.5% vs 43.8%). The held-out set is small (16 subjects), so its estimates carry wide uncertainty; the consistent direction of the r and slope lift is the load-bearing result.

**Figure 5.**
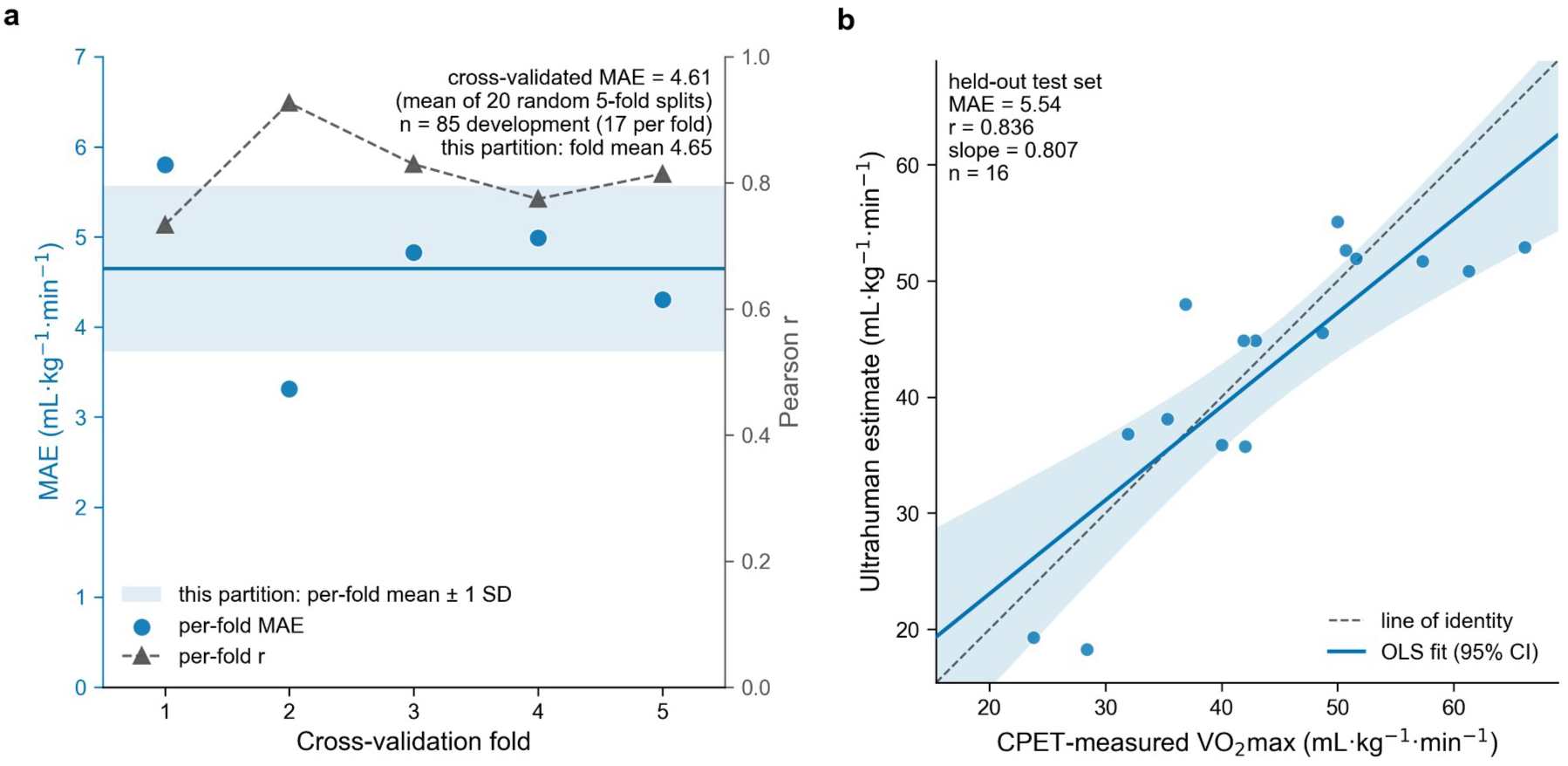
Out-of-sample generalization of the Ultrahuman estimate across methodological (random) splits. (a) Per-fold cross-validation on the 85-subject development set (5-fold scheme, 17 subjects held out per fold): per-fold mean absolute error with a mean line and ±1 SD band, and per-fold Pearson r as a second series. (b) Held-out test set agreement scatter (n=16): the Ultrahuman estimate against CPET-measured VO2max with the line of identity and the fitted regression line (r=0.836, slope=0.807, MAE=5.54). The held-out test set n is small (16), so its estimates carry wide confidence intervals; the consistent lift in r and slope on unseen subjects is the load-bearing result.

**Table 3.** Out-of-sample generalization of the Ultrahuman estimate.

| Split | n | MAE | Pearson $r$ | OLS slope | Within 5 |
| --- | --- | --- | --- | --- | --- |

| | | | | (95% CI) | $\text{mL}\cdot\text{kg}^{-1}\cdot\text{min}^{-1}$, % |
| --- | --- | --- | --- | --- | --- |
| Full-cohort analysis | 101 | 4.68 | 0.792 | 0.708 (0.581 to 0.838) | 60.4 |
| Cross-validation | 85 | 4.61 | 0.776 | 0.682 (0.544 to 0.822) | 59.9 |
| Held-out test set<sup>a</sup> | 16 | 5.54 | 0.836 | 0.807 (0.455 to 1.170) | 56.3 |
| Default configuration, everyone-medium<sup>b</sup> | 101 | 5.16 | 0.763 | 0.616 (0.495 to 0.742) | 56.4 |

MAE is in $\text{mL}\cdot\text{kg}^{-1}\cdot\text{min}^{-1}$ . $r$ is the Pearson correlation with CPET peak $\text{VO}_2$ ; slope is the ordinary-least-squares regression slope on CPET. Cross-validation is 5-fold cross-validation on the 85-subject development set (each fold holding out 17 subjects while the remainder are used to fit the calibration, rotated across all five folds). Splits are methodological (random) partitions. Slope 95% CIs are participant-level percentile bootstraps (5000 resamples, seed 42) computed with each row's evaluated predictions held fixed.
<sup>a</sup> The held-out test set of 16 subjects was excluded from the cross-validation folds and from the search keep/discard loop, and was consulted once to choose between two final configurations before evaluating the locked estimate, so it provides a near-out-of-sample (quasi-independent) read rather than a strictly untouched test set; its $n$ is small (16), so its estimates carry wide confidence intervals.
<sup>b</sup> The everyone-medium default is the default deployed (unconditioned) path: when the optional self-reported fitness level is absent, all users are scored on the medium tier. Without the fitness input the full-cohort MAE is 5.16 (vs 4.68 conditioned), and the Ultrahuman estimate still outperforms every published prediction equation on both regression slope (0.616 versus 0.317 to 0.420 for the published equations in Table 2) and Lin's CCC (0.726, 95% CI 0.633 to 0.797, versus 0.277 to 0.560).

### Default-configuration result

Because the headline assumes a self-reported fitness level, the estimate was also evaluated in everyone-medium configuration, which scores every subject on the medium tier. Full-cohort MAE was 5.16 mL·kg⁻¹·min⁻¹ (r 0.763, slope 0.616, 56.4% within 5), against 4.68 with a fitness level provided, and the same pattern held out of sample (cross-validation MAE 4.69, r 0.757, slope 0.657; held-out test MAE 6.05, r 0.788, slope 0.724, n=16; the two configurations are compared on MAE across all three splits in Figure S2). Even without the fitness input, the default configuration outperformed every published equation on both slope (0.616 versus 0.317 to 0.420) and Lin’s CCC (0.726, 95% CI 0.633 to 0.797, versus 0.277 to 0.560; Table 2).

### Population-scale behavior

To characterize deployment behavior beyond the paired corpus, each estimator was applied at population scale to the 181,133-user cohort on the default everyone-medium path. With no CPET ground truth at this scale the analysis is distributional only; no agreement metric or slope is reported here. The Ultrahuman estimate lands at a realistic level and spreads wide: population median 32.6 mL·kg⁻¹·min⁻¹ (IQR 25.4 to 39.3, width 13.9; mean 32.3), with 5.1% of users at the low-fitness floor (≤16). The published formulas read higher and narrower, with Uth at a median of 49.3 (about 17 mL·kg⁻¹·min⁻¹ above the Ultrahuman median), HUNT 42.5, Jurca 36.0, FRIEND 35.4, and Jackson 34.2, all on the same denominator; the Ultrahuman interquartile width (13.9) was about half again as wide as the widest published equation (Uth and Jackson at 9.2) and nearly double the narrowest (HUNT at 7.3), so the estimate resolves the population across a broader fitness range rather than clustering users near the mean (Figure 4c).

### Physiological and cardiometabolic correlates at population scale

A validated estimate of VO2max should rank people along a gradient of true fitness, and that gradient should be visible in physiological measurements the estimate does not use. We tested this as a construct-validity check, ranking 181,133 Ring users into fitness deciles within sex and adjusted for age, so the bottom-decile (low-fitness, n=18,112) and top-decile (high-fitness, n=18,114) groups were balanced on age (32.6 versus 33.1 years) and sex (68.9% female in both) by construction (Table 4; Figure S3). The algorithm’s inputs separated the deciles as their role requires. Night RHR was 17.4 beats·min⁻¹ lower (median 66.6 versus 49.2) and BMI 12.8 kg·m⁻² lower (median 34.2 versus 21.4) in the high-fitness group (these are Mann-Whitney median contrasts; Table 4 reports the corresponding decile means). Ring sleep measures the algorithm does not use tracked the gradient on five of the six markers examined. Relative to the low-fitness decile, the high-fitness decile had a higher sleep score, modestly longer and more efficient sleep, more rapid-eye-movement (REM) sleep, and less wakefulness after sleep onset, each separating the deciles (FDR q<.001) and retaining an association after adjustment for age, sex, and BMI. The sixth marker, deep sleep as a percentage of the night, ran the other way, being slightly lower in the high-fitness decile (Cohen’s d -0.26); deep sleep is a share of a night whose length also differs between the deciles, and we do not read it as evidence that fitter people obtain less deep sleep (per-marker values in Multimedia Appendix 1, Supplementary Table C).

**Table 4.** Cohort characteristics by validated VO2max fitness decile (population correlate analysis, N=181,133)

| Characteristic | Overall | Low-fitness (P10) | High-fitness (P90) |
| --- | --- | --- | --- |
| Users, n | 181,133 | 18,112 | 18,114 |
| Female, % | 68.9 | 68.9 | 68.9 |
| Age, y, mean (SD) | 34.8 (11.6) | 32.6 (7.8) | 33.1 (11.6) |
| BMI, kg·m<sup>-2</sup>, mean (SD) | 26.0 (5.1) | 34.6 (5.3) | 21.3 (1.8) |
| Night RHR, bpm, mean (SD) | 57.7 (7.2) | 66.6 (6.0) | 48.6 (3.3) |
| Validated VO<sub>2</sub>max, mL·kg<sup>-1</sup>·min<sup>-1</sup>, mean (SD) | 32.3 (9.4) | 19.1 (4.2) | 44.8 (5.7) |
| Daily steps, mean | 6,391 | 5,369 | 7,506 |

Fitness deciles were defined within sex on the age-adjusted residual of the validated estimate, so the low and high groups are balanced on age and sex by construction. Values are mean (SD) except n, % female, and daily steps.

The independent test came from the two channels measured by separate instruments, where shared-sensor coupling cannot explain the result. Among users with concurrent continuous glucose monitoring (Ultrahuman M1 CGM; n=2,597, of whom 399 fell in the low-fitness and 222 in the high-fitness decile), the high-fitness decile had a 13.2 mg·dL⁻¹ lower median glucose (103.7 versus 90.5), spent 18.1 percentage points more time in target range (glucose 70 to 110 mg·dL⁻¹; 61.5% versus 79.6%), and showed lower glucose variability; each association was retained after adjustment for age, sex, and BMI (adjusted β −0.41, +0.42, and −0.26 per SD of estimated VO2max; all q<.001). The venous blood panel (Ultrahuman Blood Vision) reproduced this signal and extended it across glycemic and lipid markers: the high-fitness decile had lower fasting glucose (median 91.9 versus 86.5 mg·dL⁻¹, n=7,861 assayed; adjusted β −0.19), HbA1c (5.40% versus 5.20%, Cohen’s d −0.49; β −0.12), HOMA-IR (3.55 versus 1.02, n=1,611 assayed; β −0.15), triglycerides (114 versus 69 mg·dL⁻¹, n=15,203 assayed; β −0.18), and very-low-density-lipoprotein cholesterol (24.0 versus 14.8 mg·dL⁻¹; β −0.18; all q<.001). The same direction held for HDL cholesterol (45.0 versus 60.0 mg·dL⁻¹; adjusted β +0.19), fasting insulin (15.5 versus 4.74 µIU·mL⁻¹; β −0.10), and high-sensitivity C-reactive protein (4.03 versus 0.61 mg·L⁻¹; β −0.10), each more favorable in the high-fitness decile and each retaining its association after BMI adjustment. Among the low-density-lipoprotein-burden markers, the between-decile separation of LDL cholesterol and apolipoprotein B did not survive adjustment for BMI (adjusted β +0.03 and −0.02), and that of non-HDL cholesterol attenuated to a standardized β of −0.04 (95% CI −0.07 to −0.01); their gradient is therefore carried largely by the body-mass term. Total cholesterol did not separate the deciles at all (Cohen’s d +0.01, FDR q=.89). The pre-specified negative control behaved as a true-fitness marker should: lipoprotein(a) did not differ significantly between deciles (FDR q=.37). Per-marker unadjusted effect sizes (Cohen’s d) are summarized in Figure 6, the headline decile contrasts in Figure 7, and both the unadjusted and the age-, sex-, and BMI-adjusted results, with the number of users assayed and the number in each decile, in Multimedia Appendix 1 (Supplementary Tables A–E).

**Figure 6.**
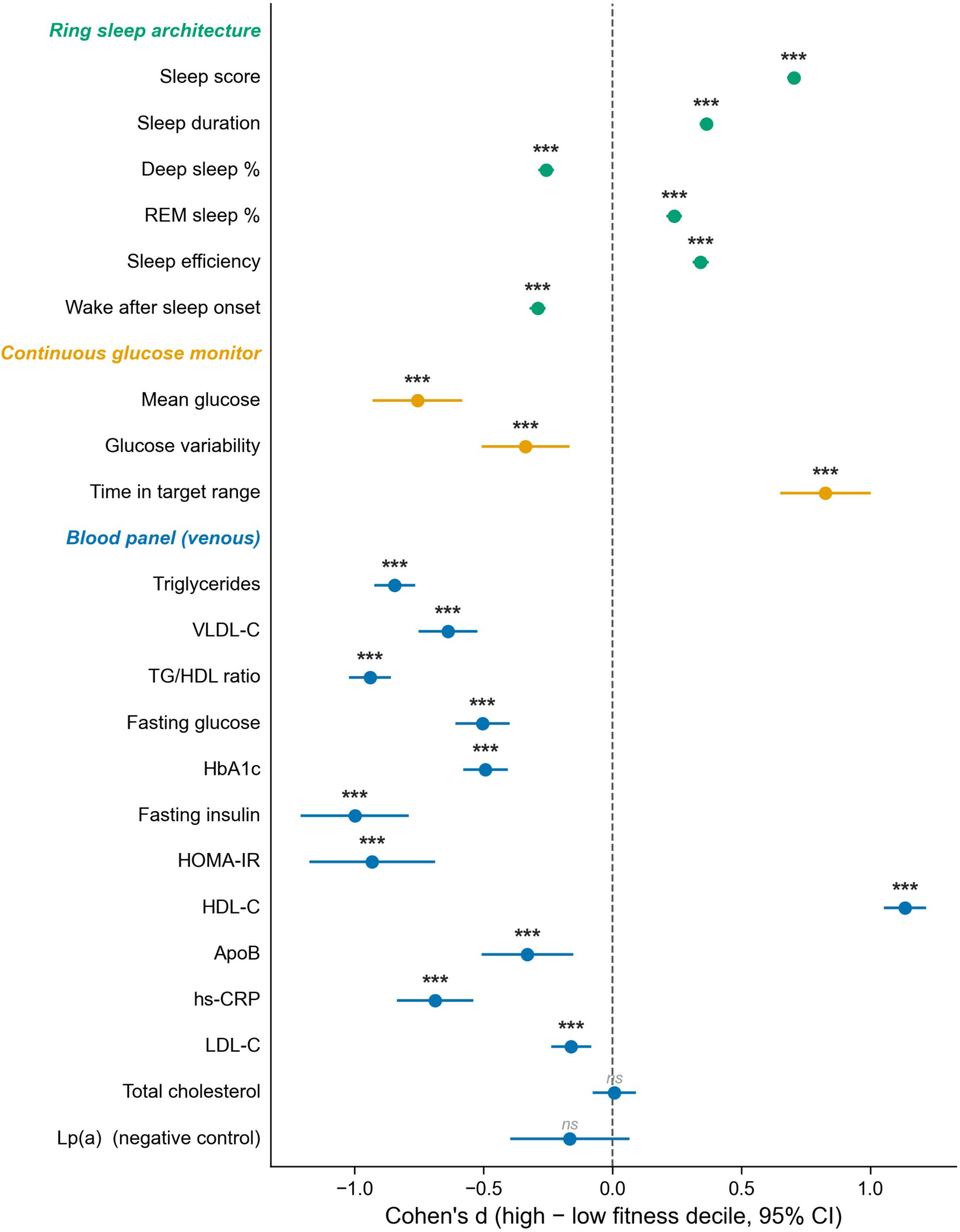
Effect size (Cohen’s d) of the high-versus low-fitness decile difference for correlates measured independently of the Ultrahuman algorithm, in the population cohort (N=181,133; deciles defined within sex and adjusted for age). Each row shows Cohen’s d with its 95% confidence interval (positive = higher in the high-fitness decile), grouped by measurement layer: the ring’s own sleep architecture, continuous glucose monitoring, and a venous blood panel. Deep sleep, expressed as a percentage of a night whose total length also differs between the deciles, is the one sleep marker that runs counter to the high-fitness decile’s otherwise more favorable sleep profile, and is not read as fitter users obtaining less deep sleep. The marker above each point is the Benjamini-Hochberg FDR-corrected Mann-Whitney significance of the between-decile difference (*** q<.001, ** q<.01, * q<.05; ns, not significant). Lipoprotein(a) is the pre-specified negative control. Ring confidence intervals are narrow because each decile contains roughly 18,000 users, whereas the CGM and blood intervals are wider owing to smaller subsamples. Age-, sex-, and BMI-adjusted associations and full per-marker numbers are given in Multimedia Appendix 1 (Supplementary Tables A–E).

**Figure 7.**
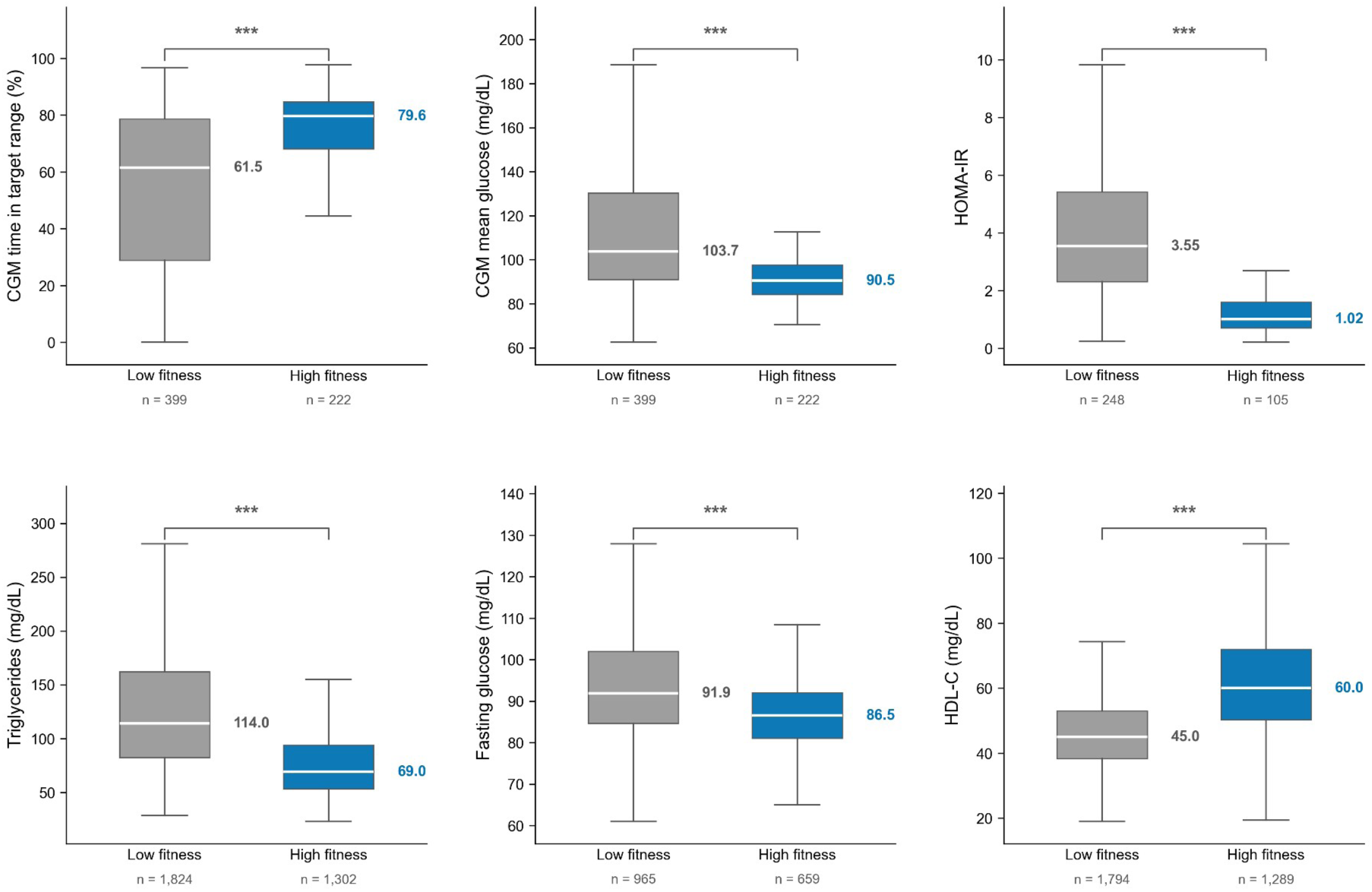
Cardiometabolic health across validated-VO2max fitness deciles, measured by instruments independent of the ring. Box plots (median, interquartile range; whiskers to 1.5×IQR, outliers omitted) for the low-fitness (bottom-decile) and high-fitness (top-decile) groups across six headline markers: continuous-glucose time in target range and mean glucose, HOMA-IR, triglycerides, fasting glucose, and HDL cholesterol. Group medians and per-group n are annotated; each y-axis spans the Tukey whisker range across both groups rather than the full data range, so the long-tailed HOMA-IR and triglyceride distributions do not compress the remaining boxes. All six contrasts are FDR-significant (q<.05). Values are unadjusted decile contrasts; all six markers shown retain an association with the estimate after adjustment for age, sex, and BMI (Multimedia Appendix 1, Supplementary Tables A–E).

## Discussion

### Principal Results

In a paired ring-CPET cohort of 101 participants, the Ultrahuman Ring AIR estimate agreed with laboratory CPET substantially better than five published non-exercise and heart-rate prediction equations, with a self-reported fitness level provided, and was best on every axis examined, including the tightest Bland-Altman limits of agreement.

A wearable estimate can correlate with CPET and still disagree with it, and that gap is what these results turn on. Each published equation either reads high at the group level or, where its mean bias is small, masks severe range compression: slopes of 0.32 to 0.42 mean the estimator moves under half a unit for each unit of true VO2max, so low-fitness participants read high and high-fitness participants read low while the cohort mean still lands close to the truth. This is the failure mode that defeats the marker’s intended use, because the mortality association of low fitness is actionable only if the estimate can identify who sits at the low end. Pearson r and group-mean MAE are both partly blind to it, which is why we lead with slope and CCC; raising the slope to 0.71, from 0.42 for the best published equation, and lifting CCC to 0.79 is a shift from estimates that rank individuals weakly to one that orders them across most of the fitness range. The small mean bias shows the ordering was recovered without trading away group-level calibration. The comparator pattern explains why the ensemble outperforms its parts: each single equation read high and range-compressed as its structure predicts, and because the components carry complementary, partly offsetting biases, an ensemble that reweights and calibrates them, with max-HR decoupled from same-day activity, can correct the shared compression no single equation corrects alone. The same signature appears at population scale, where the estimate’s landing distribution is markedly wider than any published equation’s, consistent with an estimator that preserves the fitness range rather than collapsing users toward a central value (Figure 4c).

Out-of-sample, the estimate held: on the held-out test set (n=16) slope and r rose rather than fell relative to cross-validation while MAE stayed flat. That signature, ordering preserved or improved on unseen subjects with error magnitude unchanged, is what a well-fitted ordering relationship predicts and a per-subject overfit would not, since an overfit would inflate in-sample agreement and decay out of sample. The small held-out n widens the uncertainty, and its slope interval overlaps the cross-validation interval, so the evidence is that the ordering did not decay out of sample rather than that it improved.

### Comparison with Prior Work

Consumer wearables that report VO2max share the weakness this comparison foregrounds: wrist-worn devices from the major manufacturers achieve acceptable group-level means but compress the individual range, so correlation with CPET is moderate while concordance is lower, the pattern the INTERLIVE review documents across its fourteen single-population studies [5]. Two points sharpen the comparison. First, Apple’s Cardio Fitness white paper reports a small central error (bias about 1.2 to 1.4 mL·kg⁻¹·min⁻¹, SD about 4.4 to 4.7) and a high intraclass correlation (about 0.86 to 0.89) [14], but those are the manufacturer’s own figures, and that intraclass correlation indexes longitudinal test-retest reliability, the consistency of a user’s estimate with one obtained at least four weeks earlier, rather than agreement against an independent maximal reference. The Firstbeat/Garmin white papers likewise disclose the submaximal model and the Londeree relationship while withholding the implementation [15,16]; the present study instead reports against a maximal-effort CPET reference and treats range compression as the explicit target rather than a residual to be noted. Second, by reporting CCC and slope alongside r, it makes individual-level agreement visible where a flattering mean would otherwise conceal it, a metric discipline prior wearable validations focused on r and group-mean error have largely not applied. The estimate validated here is the same VO2max input used by our previously reported wearable cardiovascular fitness-age index, which had treated that input as fixed [32]; the present study supplies its CPET validation in a separate cohort.

### Construct Validity at Population Scale

The population analysis tests a different claim from the CPET comparison: not whether the estimate matches a reference value, but whether it orders people the way true fitness does. Across 181,133 users, the high-fitness decile carried the cardiometabolic profile that laboratory work attaches to high fitness, namely lower fasting glucose and insulin resistance, lower triglycerides, higher HDL cholesterol, lower systemic inflammation, and better glycemic control on a continuous sensor [1,33,34]. Two features argue this reflects fitness rather than an artifact of construction. First, the association that carried most weight was the one a separate instrument confirmed: the agreement of a ring estimate, a subcutaneous glucose sensor, and a blood draw on the same glycemic axis is what separates criterion validity from shared-method coupling, and the glycemic and triglyceride associations survived adjustment for BMI, as did the HDL-cholesterol and inflammatory associations the literature attaches to fitness [34,35,36]; the low-density-lipoprotein-burden markers were the exception, their between-decile separation either not surviving that adjustment (LDL cholesterol, apolipoprotein B) or attenuating to a standardized β of −0.04 (non-HDL cholesterol), so their gradient tracked body mass. Second, the negative control held: lipoprotein(a), set largely by genotype and not lowered by training [31], showed no significant between-decile difference (FDR q=.37), as did total cholesterol; a spurious healthy-user gradient would have moved these too. Its age-, sex-, and BMI-adjusted continuous coefficient is small but nominally non-null (β −0.10, 95% CI −0.19 to −0.01), consistent with residual age and body-mass structure rather than a training effect; the pre-specified negative-control test is the unadjusted decile contrast (Multimedia Appendix 1, §3.4). The analysis is cross-sectional, so it describes how fit and less-fit users differ rather than the effect of changing fitness; users who carry a glucose sensor or buy a blood panel are self-selected and more health-engaged, and binning used an estimate that incorporates BMI, which is why the adjusted models, not the raw decile contrasts, carry the interpretive weight. The population analysis also relies on self-reported age, sex, height, and weight, which can contain entry errors; such errors are non-differential with respect to fitness and would attenuate the associations toward the null rather than create them.

### Limitations

We state each limitation with its mitigation. The headline accuracy assumes an optional self-reported fitness level the default path does not collect, so by default the estimate runs every user on the medium tier; without it the full-cohort MAE rises to 5.16 mL·kg⁻¹·min⁻¹ (versus 4.68), r is 0.76 and slope 0.62, and the estimate reads 5.7 mL·kg⁻¹·min⁻¹ low on average in the high-fitness band (CPET peak VO2 ≥45 mL·kg⁻¹·min⁻¹, n=44), yet still outperforms every published equation on slope and CCC (0.73 versus 0.28 to 0.56). This gap can be closed by collecting a coarse self-reported fitness level. The estimate’s slope of 0.71 remains below 1, so some range attenuation persists; this is field-universal, with wrist-worn devices showing concordance typically in the 0.5 to 0.75 band [37,38], and we report slope and CCC rather than r alone so the residual stays visible. The estimate rests primarily on night-time RHR and demographics, and resting-only signals appear to cap the achievable correlation near 0.75 to 0.79; closing the gap likely requires a sub-maximal effort channel the ring does not yet exploit, which we name as the principal accuracy-side direction for future work. Residuals are largest at the extremes, with the widest errors in the low-fitness band (Figure S4); a band-stratified error model on a larger sample is named as future work. At population scale the distribution lands at a realistic level with a wide spread and a low-fitness floor, but there is no CPET ground truth there, so we confine the validated agreement quantities to the n=101 corpus and treat the population median strictly as a calibration of level. The held-out test set (n=16) and cross-validation set (n=85) are modest, so the generalization evidence rests on the consistent direction of the r and slope lift with flat MAE rather than on a large independent sample. Finally, the validation pairs a single laboratory CPET with passively collected ring signals, so post-test baseline timing and the use of an age-predicted (Tanaka) rather than measured maximum heart rate are caveats; the max-HR decoupling removes the activity-confounding component, and in the 76 of 101 participants whose CPET record carried an achieved peak heart rate, substituting the measured peak for the age-predicted value left concordance unchanged (CCC 0.80 to 0.80), slightly increased error (MAE 4.23 to 4.57 mL·kg⁻¹·min⁻¹; paired P=.046), and raised the slope only modestly (0.69 to 0.74; change +0.04, 95% CI 0.02 to 0.07), so on this subset the age-predicted maximum heart rate accounts for at most a small part of the range compression rather than being its source. The estimate’s structure and the identity and functional form of every component are disclosed, and all five comparator formulas are published in full, but the fitted ensemble weights, calibrated coefficients, offset magnitude, activity-tier multipliers, and smoothing constant are proprietary and are not released, so the estimate cannot be reimplemented independently from this report; this follows the disclosure boundary set by comparable manufacturer reports [14,15,16]. The reference is peak oxygen uptake (VO2peak): effort was encouraged to volitional exhaustion and verified against standard indicators, but a plateau in oxygen uptake was not required and a respiratory exchange ratio of 1.10 was not reached by every participant, so the reference reflects the highest attained rather than a plateau-confirmed value, as is common in mixed-fitness cardiopulmonary exercise testing; validating the estimate against this real-world peak keeps the comparison relevant to how it is used, and because every estimator is scored against the same reference, the relative ranking of the Ultrahuman estimate and the published equations is unaffected.

The reference test was run on two modalities, and agreement differed between them. Of the 101 participants, 78 were tested on the treadmill and 21 on the cycle ergometer, with modality unrecoverable for 2. Agreement was weaker on the cycle-ergometer arm (MAE 6.52 versus 4.24 mL·kg⁻¹·min⁻¹; slope 0.627, 95% CI 0.428 to 0.883, versus 0.778, 0.633 to 0.924), and the bias reversed sign, the estimate reading 1.86 mL·kg⁻¹·min⁻¹ high against a cycle reference and 1.09 low against a treadmill one, consistent with the lower peak oxygen uptake a cycle ergometer elicits in the same person and with an estimate that is blind to the modality it is scored against. On 21 cycle tests the difference in absolute error was not statistically resolved (Welch P=.08; Mann-Whitney P=.10) and the slope intervals overlap, so this is a direction rather than an established moderator effect. Both modalities used the same metabolic cart and the ramp protocols specified above, and every estimator is scored against the same reference, so the relative ranking of the methods is unaffected; a modality-stratified analysis on a larger cycle sample is named as future work.

Two further cohort characteristics were not captured, and each bounds what could be tested. Medication use and comorbid conditions were not captured as analysis variables, so we could not test whether agents that alter resting heart rate, notably beta-blockers, degrade an estimate that rests on night-time resting heart rate and an age-predicted maximum heart rate. The cohort was also young (mean age 32.6 years, SD 7.5; range 19 to 56), with only 6 participants aged 45 years or older, so performance in older adults, in whom the age-predicted maximum-heart-rate term carries more weight, is untested. Chronotropic-medication screening and an older sample are named as requirements for the prospective external validation set identified below.

The cohort was drawn from a single site and is Indian, which raises the question of how the estimates generalize to other populations. The cohort spanned a wide fitness range (CPET peak VO2 21.6 to 66.1 mL·kg⁻¹·min⁻¹), including many high-fitness individuals, so agreement was tested across the fitness spectrum rather than only within the lower band where South Asian population means tend to sit; the high end of the range is therefore validated rather than extrapolated. The components on which the ensemble rests are grounded in physiological relationships not specific to any one population: age-predicted maximal heart rate is governed predominantly by age [20], and the heart-rate-ratio approach derives from the Fick principle and reserve ratios rather than a population-specific regression [17,39]. Because these relationships are structural, we expect them to transfer, with any between-population discrepancy expressed primarily as a shift in level. Consistent with this, direct measurement has documented lower mean fitness in South Asian than in BMI-matched European men [11], and single-population validation is itself the prevailing standard in this field [5]. Adapting the algorithm to a new population is therefore best framed as re-estimating its level rather than re-deriving its structure: miscalibration on transport is typically resolved by re-estimating an intercept (calibration-in-the-large) while leaving structure intact [40,41], and an individual-participant-data meta-analysis across nineteen countries provides a precedent in which a one-country model generalized to all others after only a per-population intercept adjustment [42]. South Asian individuals also carry higher body fat and lower resting energy expenditure at an equivalent BMI [43,44], a difference conventionally handled with population-specific offsets rather than a change of structure [45,46], and contemporary wearable VO2max models continue to be developed and validated for specific national cohorts [47,48]; this supporting evidence is detailed in Multimedia Appendix 1 (§2). We identify this transparent offset calibration, and its prospective external validation, as the principal direction for future generalizability work.

### Conclusions

Within the scope of a single-site Indian cohort of 101 participants, the Ultrahuman Ring AIR VO2max estimate agreed with laboratory CPET substantially better than five published prediction equations when a self-reported fitness level was provided, and the two-thirds lift in the regression slope relative to the best of those equations, not the modest difference in mean error, identifies range-compression correction as the source of the gain. An estimate built from resting physiology and demographics can be made to order individuals across most of the fitness range, not merely to land the cohort mean, the property the marker’s mortality association actually requires, and this was achieved by an automated search that selected the weights of an interpretable ensemble of published equations rather than by fitting an opaque model, so the improvement is auditable component by component. The estimate generalized to a held-out test set on the axes that matter (r and slope), and at population scale produced a wide distribution with a realistic low-fitness floor and ordered users along independent cardiometabolic gradients consistent with true fitness. What remains open is generalization beyond this cohort, framed as re-estimating the population level rather than re-deriving the model. Three directions follow: a multi-region CPET validation set with a transparently re-fitted population offset; an opt-in capture of a coarse self-reported fitness level to close the everyone-medium gap; and a sub-maximal effort channel to move past the resting-signal correlation ceiling.

## Supporting information

TRIPOD Reporting Checklist

STARD Reporting Checklist

## Acknowledgements

The authors thank the participants of the Ultrahuman Performance Lab VO2max cohort, who completed maximal cardiopulmonary exercise testing and wore the study ring. We thank the Performance Lab exercise-physiology staff who operated the metabolic cart, supervised the graded-exercise protocol, and curated the test records (the CPET operators and laboratory director are named in the Author Contributions).

## Funding Statement

This study was funded by Ultrahuman Healthcare Pvt Ltd, Bangalore, India, the manufacturer of the smart ring evaluated and the employer of all authors. Study design, cardiopulmonary exercise testing, data analysis, interpretation, and preparation of the manuscript were carried out by the authors in the course of their employment; no separate grant number applies. Because the funder is also the device manufacturer, and because the analysis is retrospective and was conducted without a registered protocol or a pre-registered statistical analysis plan, we state its mitigations explicitly: five independently published prediction equations were benchmarked alongside the estimate throughout, a held-out test set was withheld from the configuration search, lipoprotein(a) was pre-specified as a negative control in the population analysis, and data extraction and de-identification were completed before, and independently of, the agreement analysis. No additional external funding was received.

## Conflicts of Interest

All authors are employees of Ultrahuman Healthcare Pvt Ltd, Bangalore, India, which develops, manufactures, and commercializes the Ultrahuman Ring and the VO2max estimate evaluated in this study, and which funded the work. The calibrated coefficients of the evaluated estimate are proprietary to Ultrahuman Healthcare Pvt Ltd. The authors declare no other competing interests. Full disclosure of employment, funding, and the proprietary nature of the calibrated estimate is provided as the mitigation for industry self-validation; five independently published prediction equations are reported on every axis as external benchmarks.

## Data Availability

A de-identified per-subject table of CPET-measured peak VO2 and estimated VO2max, together with the analysis and figure-generation scripts, are available from the corresponding author on reasonable request. The estimation algorithm is described in full in the Methods as a transparent ensemble of published prediction equations; its calibrated coefficients are proprietary to Ultrahuman Healthcare Pvt Ltd and are not released. Individual-level raw wearable and laboratory records are not publicly available because of participant privacy restrictions. Aggregate statistics supporting the findings are reported in the manuscript and its Multimedia Appendices.

## Author Contributions

Contributions follow the CRediT taxonomy.

N.D. (Nihav Dhawale): formal analysis, methodology, validation, writing (review and editing). S.M. (Somaaya Mukundan): investigation (cardiopulmonary exercise testing), resources, data curation (laboratory peak VO2 records), supervision (VO2max data collection), writing (review and editing). A.A. (Ankit Agarwal): software, data curation, formal analysis, visualization, writing (review and editing). D.M. (Debasrija Mondal): investigation (ring data collection), data curation (ring-derived signals), writing (review and editing). A.S. (Aditi Shanmugam): data curation, software (data engineering), writing (review and editing). P.K. (Pavan Kumar): validation, writing (review and editing). M.M. (Mukul Mittal): conceptualization, resources, supervision, validation, writing (review and editing). V.N. (Vinayak Narasimhan): conceptualization, methodology, software, formal analysis, data curation, visualization, validation, writing (original draft), writing (review and editing), project administration, supervision. All authors reviewed and approved the final manuscript and take responsibility for the integrity of the work.

## Use of Artificial Intelligence

Generative artificial-intelligence tools (Claude, Anthropic) assisted with analysis scripting and copyediting of draft text. In addition, the estimate’s configuration (its ensemble weights, calibration, and component coefficients) was selected by an automated, agent-driven configuration search, an adaptation of the open-source autoresearch loop introduced by Karpathy [49], in which an artificial-intelligence coding agent iteratively proposed a single change, evaluated it by cross-validation on the development set, and kept or reverted it (Methods; Multimedia Appendix 1, §1.1 and Figure S5). All code was verified by the authors, and all statistical outputs and interpretations were independently reviewed and approved by the authors, who take full responsibility for the accuracy and integrity of the work. The deployed VO2max estimate is a transparent, closed-form ensemble of published physiological prediction equations and contains no learned black-box model evaluated at inference.

## Abbreviations

BMI: body mass index
CCC: concordance correlation coefficient (Lin) [27]
CGM: continuous glucose monitoring
CPET: cardiopulmonary exercise testing
EMA: exponential moving average
FDR: false discovery rate
HDL: high-density lipoprotein cholesterol
HOMA-IR: homeostatic model assessment of insulin resistance
LoA: limits of agreement (Bland-Altman) [28]
MAE: mean absolute error
MET: metabolic equivalent of task
OLS: ordinary least squares
PPG: photoplethysmography
RHR: resting heart rate
VO2max: maximal oxygen uptake
VO2peak: peak oxygen uptake.

## Notes

### Author Declarations

The study was conducted in accordance with the Declaration of Helsinki (2013 revision). The analysis of the de-identified paired CPET-and-ring dataset, and its planned publication, were reviewed and approved by the Medstar Speciality Hospital Ethics Committee, Bangalore, India (CDSCO Ethics Committee Registration No. ECR/1324/Inst/KA/2019/RR-24), by letter dated 04 July 2026. Both the paired ring-CPET analysis and the population-scale analysis of de-identified Ultrahuman platform records were approved under this review as retrospective observational research under the Indian Council of Medical Research (ICMR) National Ethical Guidelines (2017); neither analysis is a clinical trial under the New Drugs and Clinical Trials Rules (2019), so no prospective trial registration applies. Participants in the paired ring-CPET cohort had given written informed consent for laboratory testing, concurrent wearable data collection, and use of their de-identified data for research, under the Ultrahuman Performance Lab master consent form and, for those recruited through the VO2max Challenge campaign, an additional VO2max Challenge consent form. Participants whose images appear in this manuscript gave written consent for the publication of those images. Participants were not recruited for research and received no compensation for research participation; they attended the Performance Lab for a fitness assessment, and the analysis reported here is retrospective on their de-identified records. The population-scale analysis used de-identified Ultrahuman platform records across three channels, the ring, the continuous glucose monitor, and the venous blood panel, all collected under the Ultrahuman Privacy Policy and Terms of Service, which permit the analysis of de-identified grouped data for scientific research and to which users consent at onboarding and through continued use of the platform. For these retrospective components the Ethics Committee approved a waiver of additional consent under the ICMR National Ethical Guidelines (2017, Section 5.5); all records were de-identified before analysis.

