## Supplementary Information for "Accuracy of a Smart-Ring VO2max Estimate and Five Published Prediction Equations Against Cardiopulmonary Exercise Testing: Development and Validation Study With Population-Scale Analysis"

### Multimedia Appendix 1: Supplementary Methods, Tables, and Figures

This appendix holds the detailed methods moved out of the main text for length: the per-comparator input approximations applied uniformly across the five published prediction equations, the extended literature supporting the generalizability argument, the full per-layer population-correlate results (Supplementary Tables A–E), and the supplementary figures (S1–S6). The main text gives a one-sentence summary of each and points here.

#### 1. The Ultrahuman estimate and the published comparators

##### 1.1 Components of the Ultrahuman estimate

The Ultrahuman Ring AIR VO2max estimate is a transparent, weighted ensemble of three terms, each following the functional form of an independently published non-exercise or heart-rate equation and capturing a complementary pathway: a heart-rate-ratio term following the Uth relationship between the maximal-to-resting heart-rate ratio and VO2max [1]; a heart-rate-reserve term in the ACSM submaximal tradition [2]; and a non-exercise demographic regression in the Jackson tradition using age, sex, BMI, and activity [3]. Maximum heart rate is derived from the Tanaka age-prediction equation (208 − 0.7 × age) [4], deliberately decoupled from same-day activity to avoid a mobility-trap bias in which low daily movement depresses apparent achievable heart rate and inflates the estimate. The ensemble output is then adjusted by a population-level calibration offset and stabilized across days with exponential-moving-average smoothing. As stated in the main text, the input set, the identity and physiological meaning of every component, and the calibration philosophy are disclosed, whereas the ensemble weights, the calibrated coefficients and offset magnitude, the activity-tier multipliers, and the smoothing constant are proprietary.

The ensemble weights, the calibration, and the per-component coefficients were not set by hand: they were selected by an automated configuration search, an adaptation of the open-source "autoresearch" loop introduced by Karpathy [5]. In the original autoresearch, an automated coding agent repeatedly edits a model's training code, runs a short fixed-budget experiment, and keeps or reverts each edit according to whether a held-out validation metric improves. We retained that commit-or-revert hill-climbing loop but adapted the candidate, the evaluator, and the objective to our problem: the candidate is the configuration of our published-equation ensemble (rather than a neural-network training script), the evaluator is a cross-validated agreement criterion against CPET on the development set (rather than a single-run validation loss), and the objective rewards individual-level agreement, not just average error. The overall process is summarized in Figure S5. Starting from a baseline configuration, the search ran a fixed budget of 500 candidate experiments; at each step it proposed one change to the configuration (the ensemble weights, the calibration, the per-component coefficients, the activity-tier multipliers, or optional structural corrections), scored the candidate, and kept the change only when it lowered the search objective and passed pre-set quality gates, otherwise reverting to the current best. The objective was a composite cost computed on the 85-subject development set, dominated by repeated, seed-averaged 5-fold cross-validated mean absolute error (its single largest term), with smaller terms rewarding higher correlation, a regression slope near 1, low stratum-specific bias, and stability across folds and seeds, a small penalty computed on a separate four-subject holdout outside the study cohort, and a parameter-count penalty, among other minor terms. This composite was used so that the search optimized individual-level agreement (slope and concordance) rather than group-mean error alone. The 16-subject held-out test set was excluded from the cross-validation folds and from the search keep/discard objective. After the search, it was consulted once to choose between the final candidate configuration and one alternate, and was then used to evaluate the locked estimate; it therefore provides a near-out-of-sample (quasi-independent) read rather than a strictly untouched test set. Because the repeated cross-validated error was itself the search objective, the development-set cross-validation figures reflect in-development optimization and are not an independent estimate of generalization. The locked estimate's level calibration was fitted on the full corpus, which is a further reason the full-cohort row is an in-sample fit; the cross-validation and held-out rows refit that calibration within training data only. The resulting estimate is the weighted ensemble described above (a heart-rate-ratio term, a heart-rate-reserve term, and a non-exercise demographic term, evaluated on a Tanaka age-predicted maximum heart rate); the calibrated weights, coefficients, calibration offset, activity-tier multipliers, and smoothing constant remain proprietary. The locked estimate's agreement on the 16-subject held-out test set, reported alongside the development-set cross-validation, is given in the main text (Table 3 and Figure 5).

The search's progress is shown in Figure S6. Over the 500 experiments, the running-best composite cost fell monotonically from the seed configuration (3.7534) to the locked configuration (3.4002), a 9.4% reduction, through 41 committed changes; the remaining experiments were discarded, rejected by a quality gate, or errored without usable metrics. The improvement was concentrated on range agreement rather than mean error: along the committed-best trajectory the cross-validated regression slope rose from 0.633 to 0.692 and the cross-validated correlation from 0.730 to 0.775, while the cross-validated MAE changed little, and non-monotonically, from 4.72 to 4.50 $mL\cdot kg^{-1}\cdot min^{-1}$. This is the property the present validation argues matters (slope and concordance, not group-mean error), and it is why the search minimized a composite criterion rather than MAE alone. Because that criterion weights several terms, the accuracy-versus-range-agreement plane in Figure S6b is a projection of the objective rather than a Pareto front, and the selected configuration is the composite-cost minimum, not a two-objective optimum. The values in Figure S6 are the search-time metrics on the development corpus used during the loop; the agreement statistics reported in the main text are recomputed on the refreshed cohort (Table 3), so absolute values differ slightly.

##### 1.2 Comparator equations and input approximations

Each published equation was computed on the same cohort and received the same ring-derived inputs as the Ultrahuman estimate, so the benchmark is on equal footing; every input approximation is disclosed here and carried uniformly across methods.

Where a formula was originally specified with awake or clinic resting heart rate, we supplied the ring's night-time sleeping RHR, which sits a few $beats\cdot min^{-1}$ lower; this substitution is applied uniformly to every RHR-using method and noted as a limitation. Maximal heart rate, where a comparator equation required it, was estimated from age and activity tier following the published-baseline comparator convention: low tier $220 - age$ (Fox) [6], medium tier $207 - 0.7 \times age$

(Gellish) [7], and high tier 211 − 0.64 × age (Nes) [8]. The Ultrahuman estimate itself does not use this tier-varying rule; as described in §1.1 it takes a single Tanaka age-predicted maximum heart rate (208 − 0.7 × age), deliberately decoupled from same-day activity. The activity tier (low/medium/high) was derived from the 7-day step average using cut-points of fewer than 5000, 5000 to 10,000, and more than 10,000 steps. For non-binary sex, equations used the midpoint of the male and female codings. Height was missing for 2 of the 101 participants, one never recorded and one set to missing by the plausibility screen (Methods; §1.3). None of the published equations specifies a rule for a missing height, so for these two the sex-specific cohort median height was imputed (173.75 cm; both are male). The median is taken over the participants with a usable height, so the erroneous value does not enter it.

The five published comparators were:

- *Uth et al. (heart-rate-ratio method)* [1]: VO2max = 15.3 × (HRmax / HRrest), with HRmax estimated as above and HRrest the night RHR.
- *Jurca et al. (non-exercise CRF model, NASA equation)* [9]: CRF (METs) = 18.07 + 2.77 × G − 0.10 × age − 0.17 × BMI − 0.03 × RHR + SR-PA, with G = 1 (male), 0 (female), 0.5 (other), and VO2max = CRF × 3.5 $mL \cdot kg^{-1} \cdot min^{-1}$. We used the NASA variant (the recommended, lowest standard-error-of-estimate model). The same disclosed approximations as the other comparators were applied: the ring's night-time sleeping RHR was supplied as the resting heart rate, and the activity tier was mapped to the model's self-reported physical-activity (SR-PA) score on the 5-point NASA scale, using the interior levels (low → Level 2, weight 0.32; medium → Level 3, 1.06; high → Level 4, 1.76) and avoiding the extreme inactive (Level 1, 0.00) and very-active (Level 5, 3.03) anchors, because the ring infers activity rather than administering the questionnaire.
- *Jackson et al. (non-exercise BMI regression)* [3]: VO2max = 56.363 + 1.921 × PA-R − 0.381 × age − 0.754 × BMI + 10.987 × G, with G = 1 (male), 0 (female), 0.5 (other), and the self-reported physical-activity rating PA-R (0 to 7 scale) mapped from the activity tier as low → 2, medium → 4, high → 6 (a documented approximation, because the ring infers activity rather than administering the PA-R questionnaire). We used the BMI variant because the ring provides BMI rather than body composition.
- *FRIEND registry reference equation (de Souza e Silva et al.)* [10]: VO2max = 45.2 − 0.35 × age − 10.9 × S − 0.15 × weight_lb + 0.68 × height_in − 0.46 × mode, with S = 1 (male), 2 (female), 1.5 (other); weight and height were converted to pounds and inches. Treadmill mode (mode = 1) was applied uniformly to every subject as a disclosed approximation, including the 21 participants tested on a cycle ergometer (Methods). Because mode enters as a constant −0.46 × mode, a cycle coding would lower the FRIEND estimate for those 21 by 0.46 $mL \cdot kg^{-1} \cdot min^{-1}$ and would not change its ranking of individuals; recomputing FRIEND with the recovered per-participant modality moves its full-cohort MAE from 6.34 to 6.35 $mL \cdot kg^{-1} \cdot min^{-1}$ and its slope from 0.392 to 0.393, so the comparison is unaffected on the slope and concordance axes the benchmark turns on. FRIEND already reads low on this cohort (bias −1.20), so the uniform treadmill coding is the more generous of the two codings for the comparator, consistent with the convention applied to the other input approximations.
- *HUNT/Nes et al. (non-exercise VO2peak model)* [11]: sex-specific regressions on age, waist circumference, RHR, and a physical-activity index. Because the ring does not measure waist

circumference, it was approximated from BMI (waist ≈ 2.5 × BMI + 25 for men, + 17 for women, + 21 for other), and the HUNT physical-activity index (0 to 15 scale) was mapped from the activity tier as low → 2, medium → 7, high → 12. Requiring two inputs a wearable does not capture (waist and a structured activity questionnaire), HUNT is the most approximated comparator.

These input approximations are a deliberate, conservative design choice rather than a handicap on the comparators, and we note here the questions a reader may reasonably raise about the fairness of the benchmark. Each published equation was originally derived with at least one input a passively worn ring does not measure directly (clinic resting heart rate, a structured physical-activity questionnaire, or waist circumference); supplying the ring's own approximations of these inputs is how each equation would have to be deployed on ring data, so the comparison reflects realistic wearable use rather than an artificial penalty. The same input approximations (the ring's night-time RHR in place of clinic RHR, and the activity-tier mappings) were applied uniformly across the five published equations, so no comparator is disadvantaged relative to another; where the Ultrahuman estimate itself differs from the comparators, namely in its Tanaka age-predicted maximum heart rate decoupled from activity (§1.1) and the shipped missing-height path described below, that difference is disclosed rather than hidden. Missing height is the single input the methods do not share a rule for, and the choice of rule is immaterial. The published equations have no missing-height provision, so each was supplied the sex-specific cohort median (173.75 cm), whereas the Ultrahuman estimate falls back to the rule it ships with, a neutral BMI with the stature term zeroed. Supplying the Ultrahuman estimate with the same imputed median instead moves its full-cohort MAE from 4.68 to 4.69 mL·kg$^{-1}$·min$^{-1}$, and restricting the analysis to the 99 participants with a usable height gives 4.64; the margin over the best published equation is 1.52, 1.51, and 1.49 mL·kg$^{-1}$·min$^{-1}$ under the three treatments, and the ranking of the methods is identical in each (§1.3). Where an approximation of an activity input could plausibly favor a comparator, the more generous option was taken: interior activity levels were used rather than the extreme questionnaire anchors. The night-time sleeping resting heart rate supplied wherever a resting heart rate was required sits a few beats·min$^{-1}$ below clinic resting heart rate, which raises a heart-rate-ratio estimate; because the Uth heart-rate-ratio method already over-reads on this cohort (bias +9.90 mL·kg$^{-1}$·min$^{-1}$, Table 2), that shift moves it further from the reference rather than toward it. The comparison does not rest on any such level choice. Range compression is measured by the regression slope: for the four linear comparators a uniform shift in the assumed resting heart rate produces at most a constant additive change in the estimate and so leaves the slope unchanged (their slopes of 0.38 to 0.42), while for the Uth ratio the same shift rescales the estimate by only a few percent (slope 0.32); the compression is thus a property of the equations and not of the inputs supplied. A uniform level shift instead moves the mean bias and, with it, Lin's CCC, and as noted it moves them against the over-reading heart-rate methods rather than for them. The benchmark therefore tests whether the published equations, given the best inputs a ring can provide, recover individual-level fitness; the range compression they show is a property of the equations, not an artifact of the input approximations.

##### 1.3 Anthropometric data quality and sensitivity analysis

Height and weight are self-entered profile values, so they carry data-entry error. Every screened record was therefore checked against an adult plausibility bound for height of 140 to 210 cm. The bound is not specific to this analysis: it is the same height window the 181,133-user population cohort is constructed with, so applying it to the paired cohort makes the two arms of the study consistent rather than introducing a new criterion. It was applied without reference to the CPET result or to any estimator's error, so it cannot select for participants that a given method happens to predict poorly. The bound is also far from any real measurement here: the shortest genuine height in the cohort is 150.0 cm, a clear 10 cm above the lower bound, so no true stature is at risk of being caught by it.

The screen flagged one record: a height of 127 cm recorded at a weight of 69 kg, implying a BMI of 42.8 kg·m$^{-2}$. That height is 23 cm shorter than the shortest genuine participant and is not a possible adult stature. The height and the BMI derived from it were set to missing, and the participant was retained on all remaining inputs. Weight was screened on the same basis and no value in the cohort was implausible (range 44.9 to 110.2 kg); this participant's 69 kg is unremarkable, and it is the mass the laboratory used to normalize his CPET measurement, so only the stature entry is in question, not the record as a whole.

The erroneous value was consequential because it was consumed as an input, not merely reported. At 127 cm the implied BMI of 42.8 kg·m$^{-2}$ entered every BMI-dependent comparator, and for HUNT it implied a waist circumference of 132 cm. The participant's measured CPET peak VO2 was 56.7 mL·kg$^{-1}$·min$^{-1}$, near the top of the cohort, so the published equations were being asked to reconcile a high measured fitness with an obese body habitus that did not exist, and they under-estimated him by 23 to 32 mL·kg$^{-1}$·min$^{-1}$ (Jackson 31.9, Jurca 26.0, FRIEND 24.6, HUNT 22.9); correcting the stature raised their estimates for him by 12 to 18 mL·kg$^{-1}$·min$^{-1}$. The Ultrahuman estimate was exposed to the same entry but only partly, because its shipped input-hygiene layer bounds an out-of-range height before use; correcting the input moved its estimate for this participant from 43.72 to 46.74 mL·kg$^{-1}$·min$^{-1}$ against a measured 56.7, so he remains a substantial under-estimate either way. Because the comparators were fully exposed and the Ultrahuman estimate only partly, the correction improves the comparators more than it improves the Ultrahuman estimate.

The table below reports the full agreement set under both treatments: the analysis as reported, and a sensitivity analysis in which the erroneous 127 cm is retained and every method is recomputed on it.

| Method | MAE, value retained | MAE, as reported | Pearson r, retained | r, as reported | CCC, retained | CCC, as reported |
|---|---|---|---|---|---|---|
| Ultrahuman | 4.71 | 4.68 | 0.788 | 0.792 | 0.782 | 0.787 |
| Jurca 2005 | 6.32 | 6.20 | 0.605 | 0.645 | 0.530 | 0.560 |
| FRIEND 2018 | 6.46 | 6.34 | 0.596 | 0.631 | 0.530 | 0.560 |
| Jackson 1990 | 6.86 | 6.72 | 0.569 | 0.615 | 0.513 | 0.552 |
| HUNT/Nes 2011 | 7.38 | 7.20 | 0.626 | 0.687 | 0.445 | 0.474 |
| Uth 2004 | 10.55 | 10.55 | 0.612 | 0.612 | 0.277 | 0.277 |

No conclusion depends on the choice. Every method improves or is unchanged, the ranking is identical under both treatments, and the correction is the more conservative of the two for the Ultrahuman estimate: its MAE falls by 0.03 mL·kg$^{-1}$·min$^{-1}$ while the four BMI-dependent comparators fall by 0.12 to 0.18, so the margin between the Ultrahuman estimate and the best published equation narrows from 1.60 to 1.52 mL·kg$^{-1}$·min$^{-1}$. Uth is unaffected because it is a heart-rate-ratio method and takes no anthropometric input. The held-out test set is unchanged in every respect, since the participant is in the development split.

A third treatment was available and was not taken: excluding the participant altogether. It would have reported a lower Ultrahuman MAE than the analysis above (4.63 on n=100, against the 4.68 reported on n=101), so retaining him is the more conservative option as well as the one that preserves the sample. The record contains one unusable field, not an unusable participant, and dropping a participant for a mistyped digit would discard a valid CPET measurement and a valid set of ring signals.

#### 2. Extended generalizability evidence

The main-text generalizability subsection states the argument and cites its anchor references; the supporting literature is collected here.

*Structural physiology.* The proportional relationship between the fraction of heart-rate reserve and the fraction of oxygen-uptake reserve has been examined directly, with the two fractions differing under graded testing by an approximately constant offset rather than in functional form, although that proportionality is reported to hold less cleanly during prolonged isocaloric exercise [12].

*Ethnic differences as level shifts.* South-Asian individuals carry a higher proportion of body fat and exhibit lower resting energy expenditure than European individuals at an equivalent body-mass index [13,14]. The established response to such shifts has been to retain the underlying relationship and adjust population-specific thresholds or offsets, as for ethnicity-specific adiposity and diabetes-risk cut-points [15,16].

*Single-population validation as field standard.* Single-population validation is the prevailing standard in this field: a systematic review and meta-analysis of consumer-wearable VO2max estimation identified only fourteen validation studies, each in a single population, and concluded that exercise-based estimation is accurate at the population level while individual-level error remains substantial [17]. Contemporary wearable VO2max models continue to be developed and validated for specific national populations, including Korean [18] and Chinese [19] cohorts, and flagship consumer devices have themselves been validated in small single-site samples [20].

*Re-estimating level, not re-deriving structure.* In the prediction-modelling literature, transporting a model to a new setting routinely separates discrimination, which reflects structure, from calibration, which reflects level; miscalibration on transport is typically resolved by re-estimating an intercept (calibration-in-the-large) while leaving structure intact [21,22]. A recent individual-participant-data meta-analysis across nineteen countries provides a concrete precedent: a model developed in one country generalized well to all others, and its performance improved simply by re-estimating the intercept for each population, without any change to the model's form [23].

#### 3. Population correlate analysis

This appendix details the construct-validity analysis summarized in the main text (Results, Physiological and cardiometabolic correlates at population scale), and reports the full per-layer results behind Figures 6, 7, and S3 (Supplementary Tables A–E below).

##### 3.1 Cohort and recomputed estimate

The cohort comprised de-identified Ultrahuman Ring AIR users with at least 20 nights of valid night-time resting heart rate in a fixed recent 90-day window and complete age, sex, height, and weight, yielding 181,133 users (124,886 female, 56,247 male; mean age 34.8 years, SD 11.6; mean BMI 26.0 $kg \cdot m^{-2}$, SD 5.1). User-level records were de-identified before analysis. For each user the validated estimate was recomputed with the same implementation evaluated against CPET, applied to per-user mean inputs (night-time resting heart rate, age, sex, BMI, and a step-derived activity tier) on the default medium-tier path, because no self-reported fitness level is collected at scale. The estimate was computed as a single-shot point value without the deployed exponential-moving-average smoothing, since per-user means rather than daily series were used; the smoothing affects day-to-day stability, not the cross-sectional ranking used here. The recomputed estimate had a population median of 32.6 $mL \cdot kg^{-1} \cdot min^{-1}$ (male 35.6, female 31.3), matching the population median reported in the main text (Results, Population-scale behavior).

##### 3.2 Binning, matching, and statistics

Because the estimate is sex- and age-dependent, users were ranked on the within-sex residual of the estimate after regressing it on age, and the bottom and top deciles of that residual defined the low-fitness and high-fitness groups. This within-sex, age-adjusted ranking balances the two groups on age (means 32.6 versus 33.1 years) and sex (68.9% female in both) without a separate matching step, which is the design analogue of the age and sex matching used in extreme-group fitness comparisons. Each correlate was compared between deciles with the two-sided Mann-Whitney U test, reporting the median difference with a 1000-sample bootstrap 95% confidence interval and the rank-biserial effect size, and the within-sex Spearman correlation of the marker with the continuous estimate. The adjusted association was estimated by ordinary-least-squares regression of the standardized marker on the standardized estimate with age, sex, and BMI as covariates, reporting the standardized coefficient with its 95% confidence interval; adjusting for BMI is conservative because BMI is itself an input to the estimate, so the adjusted coefficient isolates the association not carried by the body-mass term. P values were corrected within each correlate layer by the Benjamini-Hochberg procedure (FDR). Continuous glucose summaries were available for 2,597 users and venous blood markers for up to 15,203 users per marker. Because the Blood Vision panel aggregates results across laboratories, a minority of venous values were recorded in an alternate unit (lipids and glucose in $mmol \cdot L^{-1}$ rather than $mg \cdot dL^{-1}$, HbA1c in $mmol \cdot mol^{-1}$ rather than %, and the apolipoproteins in $g \cdot L^{-1}$ rather than $mg \cdot dL^{-1}$); each marker was harmonized to its conventional unit before analysis by converting values that were implausible in the conventional unit but physiological after the standard conversion, and values implausible in any unit (for example, negative entries) were discarded. HOMA-IR was computed from the harmonized components as fasting glucose ($mg \cdot dL^{-1}$) × fasting insulin ($\mu IU \cdot mL^{-1}$) / 405, and the triglyceride-to-HDL ratio likewise from the harmonized panel.

##### 3.3 Stability of decile membership across estimators

Because fitness can be estimated in several ways from the same ring inputs, the stability of decile membership across estimators was checked. The validated (recomputed) estimate and an alternative ring-based estimator ranked users concordantly (Spearman $\rho = 0.80$ over the full cohort). A transparent equal-weight ensemble of the published component equations (Uth heart-rate ratio and Jackson non-exercise, with Tanaka maximum heart rate) ranked users concordantly with that alternative estimator (ρ = 0.87) and shared about 79% of the high-fitness decile and 56% of the low-fitness decile; the heart-rate-ratio component alone agreed on more than 89% of both deciles. The high-fitness decile is therefore stable to the choice of estimator, and the low-fitness decile is more estimator-sensitive because the demographic and body-mass terms carry more weight at the low end. The correlate results are reported on the validated (recomputed) estimate.

##### 3.4 Negative control

Lipoprotein(a) was pre-specified as a negative control. Its serum concentration is largely genetically determined and is not lowered by habitual physical activity or aerobic training [24], so a true marker of cardiorespiratory fitness should show no association with it. A spurious healthy-user gradient, in which more health-engaged users record more favorable values on every assay, would instead move lipoprotein(a) along with the other markers; the flatness of its between-decile contrast (FDR q=.37; Cohen's d −0.17) is therefore evidence that the gradient reflects fitness-linked physiology rather than general health engagement. The pre-specified negative-control test is this unadjusted decile contrast; its age-, sex-, and BMI-adjusted continuous coefficient is small but nominally non-null (β −0.10, 95% CI −0.19 to −0.01), consistent with residual age- and body-mass structure rather than a training effect.

##### 3.5 Full per-layer results

The tables below report every correlate examined, by independence layer. In the n column the first number is the users with a value for that marker, which is the sample the adjusted beta and the within-sex Spearman are computed on, and the two numbers in parentheses are the low- and high-fitness decile members with a value, which are the samples the medians, IQRs, Cohen's d, and FDR q are computed on. The effect size is Cohen's d for the high- versus low-fitness decile difference (positive = higher in the high-fitness decile); the adjusted beta is the age-, sex-, and BMI-adjusted standardized association of the marker with the validated estimate; the within-sex Spearman is the rank correlation of the marker with the estimate; FDR q is the Benjamini-Hochberg-corrected Mann-Whitney p-value. "<.001" denotes a q-value below .001.

**Supplementary Table A. Dependent layer (algorithm inputs; positive controls).**

| Correlate | n (low/high decile) | Low median (IQR) | High median (IQR) | Cohen's d (95% CI) | Adjusted beta (95% CI) | Within-sex Spearman | FDR q |
|---|---|---|---|---|---|---|---|
| Night RHR (bpm) | 181,133 (18,112/18,114) | 66.6 (62.5-70.5) | 49.2 (46.9-50.9) | -3.84 (-3.87 to -3.80) | -1.56 (-1.56 to -1.55) | -0.70 | <.001 |
| BMI ($kg \cdot m^{-2}$) | 181,133 (18,112/18,114) | 34.2 (30.9-38.0) | 21.4 (20.1-22.6) | -3.40 (-3.43 to -3.37) | -0.89 (-0.89 to -0.89) | -0.64 | <.001 |
| Daily steps | 181,133 (18,112/18,114) | 5,069 (3,879-6,554) | 7,135 (5,329-9,270) | +0.80 (+0.77 to +0.82) | +0.33 (+0.33 to +0.34) | +0.19 | <.001 |

**Supplementary Table B. Autonomic layer (photoplethysmography-derived; shares the ring's sensor).**

| Correlate | n (low/high decile) | Low median (IQR) | High median (IQR) | Cohen's d (95% CI) | Adjusted beta (95% CI) | Within-sex Spearman | FDR q |
|---|---|---|---|---|---|---|---|
| Sleep HRV (ms) | 181,092 (18,107/18,110) | 45.7 (38.7-54.3) | 70.2 (60.3-81.3) | +1.62 (+1.60 to +1.64) | +1.01 (+1.01 to +1.02) | +0.58 | <.001 |

Sleep heart-rate variability shares the ring's photoplethysmography sensor and is physiologically coupled to resting heart rate, so its large association is reported here as a shared-sensor reference rather than as an independent correlate.

**Supplementary Table C. Ring sleep architecture (not used by the algorithm).**

| Correlate | n (low/high decile) | Low median (IQR) | High median (IQR) | Cohen's d (95% CI) | Adjusted beta (95% CI) | Within-sex Spearman | FDR q |
|---|---|---|---|---|---|---|---|
| Sleep score | 181,133 (18,112/18,114) | 75.3 (70.2-79.4) | 80.0 (75.6-83.6) | +0.70 (+0.68 to +0.72) | +0.33 (+0.32 to +0.33) | +0.20 | <.001 |
| Sleep duration (h) | 181,133 (18,112/18,114) | 7.03 (6.4-7.6) | 7.33 (6.8-7.8) | +0.36 (+0.34 to +0.38) | +0.09 (+0.09 to +0.10) | +0.16 | <.001 |
| Deep sleep (%) | 119,536 (11,525/11,970) | 19.0 (16.0-22.7) | 18.0 (15.0-21.0) | -0.26 (-0.28 to -0.23) | -0.12 (-0.13 to -0.11) | +0.01 | <.001 |
| REM sleep (%) | 119,560 (11,529/11,972) | 22.1 (18.3-25.4) | 23.3 (20.0-26.1) | +0.24 (+0.21 to +0.27) | +0.15 (+0.14 to +0.16) | +0.14 | <.001 |
| Sleep efficiency (%) | 119,589 (11,538/11,972) | 91.7 (88.3-94.0) | 93.3 (90.3-95.3) | +0.34 (+0.32 to +0.37) | +0.08 (+0.07 to +0.09) | +0.21 | <.001 |
| Wake after sleep onset (min) | 119,579 (11,538/11,972) | 35.0 (23.3-52.5) | 28.3 (20.0-43.3) | -0.29 (-0.32 to -0.26) | -0.06 (-0.07 to -0.05) | -0.19 | <.001 |

Sleep score is Ultrahuman's proprietary nightly sleep-quality composite (0 to 100); because its internal derivation is undisclosed and may incorporate autonomic (heart-rate-derived) features, it is reported here as a convenience summary rather than a strictly sensor-independent correlate, and the transparent architecture markers (sleep duration, efficiency, REM percentage, and wake after sleep onset) carry the layer's evidence.

**Supplementary Table D. CGM layer (continuous glucose monitor; separate sensor).**

| Correlate | n (low/high decile) | Low median (IQR) | High median (IQR) | Cohen's d (95% CI) | Adjusted beta (95% CI) | Within-sex Spearman | FDR q |
|---|---|---|---|---|---|---|---|
| CGM mean glucose ($mg \cdot dL^{-1}$) | 2,597 (399/222) | 103.7 (91.0-130.3) | 90.5 (84.3-97.6) | -0.76 (-0.93 to -0.59) | -0.41 (-0.47 to -0.35) | -0.35 | <.001 |
| CGM glucose variability | 2,597 (399/222) | 15.5 (12.8-19.1) | 14.2 (12.4-16.5) | -0.34 (-0.50 to -0.17) | -0.26 (-0.33 to -0.20) | -0.14 | <.001 |
| CGM time in target range (%) | 2,597 (399/222) | 61.5 (28.8-78.6) | 79.6 (68.2-84.7) | +0.83 (+0.66 to +1.00) | +0.42 (+0.36 to +0.48) | +0.29 | <.001 |

CGM time in target range is the percentage of continuous-glucose readings within the Ultrahuman platform's restricted metabolic-health target band of 70 to 110 $mg \cdot dL^{-1}$.

**Supplementary Table E. Blood layer (Ultrahuman Blood Vision venous panel; reference instrument).**

| Correlate | n (low/high decile) | Low median (IQR) | High median (IQR) | Cohen's d (95% CI) | Adjusted beta (95% CI) | Within-sex Spearman | FDR q |
|---|---|---|---|---|---|---|---|
| Triglycerides ($mg \cdot dL^{-1}$) | 15,203 (1,824/1,302) | 114.0 (82.4-162.0) | 69.0 (53.1-94.0) | -0.84 (-0.92 to -0.77) | -0.18 (-0.21 to -0.16) | -0.29 | <.001 |
| VLDL-C | 7,006 | 24.0 (17.0- | 14.8 (11.0- | -0.64 (-0.75 | -0.18 (-0.22 | -0.28 | <.001 |

| | | | | | | | |
|---|---|---|---|---|---|---|---|
| (mg·dL$^{-1}$) | (937/533) | 34.0) | 21.4) | to -0.53) | to -0.14) | | |
| TG/HDL ratio | 14,641 (1,771/1,255) | 2.48 (1.7-3.9) | 1.13 (0.8-1.7) | -0.94 (-1.02 to -0.86) | -0.21 (-0.23 to -0.18) | -0.33 | <.001 |
| Fasting glucose (mg·dL$^{-1}$) | 7,861 (965/659) | 91.9 (84.7-102.0) | 86.5 (81.0-92.0) | -0.50 (-0.60 to -0.40) | -0.19 (-0.23 to -0.15) | -0.25 | <.001 |
| HbA1c (%) | 12,068 (1,633/950) | 5.40 (5.2-5.8) | 5.20 (5.0-5.5) | -0.49 (-0.57 to -0.41) | -0.12 (-0.15 to -0.09) | -0.25 | <.001 |
| Fasting insulin (µIU·mL$^{-1}$) | 2,153 (302/154) | 15.5 (10.0-22.9) | 4.74 (3.2-7.0) | -1.00 (-1.20 to -0.80) | -0.10 (-0.16 to -0.03) | -0.38 | <.001 |
| HOMA-IR | 1,611 (248/105) | 3.55 (2.3-5.4) | 1.02 (0.7-1.6) | -0.93 (-1.17 to -0.69) | -0.15 (-0.23 to -0.07) | -0.40 | <.001 |
| HDL-C (mg·dL$^{-1}$) | 14,911 (1,794/1,289) | 45.0 (38.3-53.0) | 60.0 (50.3-72.0) | +1.13 (+1.06 to +1.21) | +0.19 (+0.16 to +0.21) | +0.25 | <.001 |
| ApoB (mg·dL$^{-1}$) | 2,579 (296/234) | 97.0 (81.4-113.0) | 89.0 (72.7-103.0) | -0.33 (-0.50 to -0.16) | -0.02 (-0.08 to +0.05) | -0.11 | <.001 |
| hs-CRP (mg·L$^{-1}$) | 3,917 (482/342) | 4.03 (1.9-8.1) | 0.61 (0.3-1.3) | -0.69 (-0.83 to -0.55) | -0.10 (-0.16 to -0.05) | -0.33 | <.001 |
| Homocysteine (µmol·L$^{-1}$) | 2,242 (256/209) | 12.2 (9.3-17.2) | 10.1 (7.9-15.0) | -0.37 (-0.55 to -0.18) | -0.16 (-0.22 to -0.09) | -0.10 | <.001 |
| LDL-C (mg·dL$^{-1}$) | 14,697 (1,778/1,260) | 112.1 (92.0-136.0) | 106.0 (86.9-128.0) | -0.16 (-0.23 to -0.09) | +0.03 (-0.00 to +0.06) | -0.08 | <.001 |
| Total cholesterol (mg·dL$^{-1}$) | 12,265 (1,512/1,039) | 183.0 (160.0-209.0) | 182.0 (161.0-208.4) | +0.01 (-0.07 to +0.09) | +0.05 (+0.02 to +0.08) | -0.05 | .89 |
| Non-HDL-C (mg·dL$^{-1}$) | 9,954 (1,295/802) | 136.9 (114.0-164.5) | 121.0 (99.4-143.0) | -0.42 (-0.51 to -0.33) | -0.04 (-0.07 to -0.01) | -0.15 | <.001 |
| ApoA1 (mg·dL$^{-1}$) | 1,756 (214/146) | 118.1 (108.2-133.0) | 131.0 (116.7-149.0) | +0.55 (+0.33 to +0.76) | +0.09 (+0.02 to +0.17) | +0.06 | <.001 |
| LDL/HDL ratio | 4,132 (547/318) | 2.70 (2.0-3.4) | 2.00 (1.5-2.6) | -0.74 (-0.89 to -0.60) | -0.10 (-0.15 to -0.05) | -0.19 | <.001 |
| Lipoprotein(a) (mg·dL$^{-1}$) [negative control] | 1,466 (186/128) | 17.4 (7.8-39.7) | 15.4 (6.7-31.7) | -0.17 (-0.39 to +0.06) | -0.10 (-0.19 to -0.01) | -0.03 | .37 |

In the blood layer, total cholesterol is the only marker not reaching FDR q<.05 besides the negative control. Most markers retained an association with the estimate after age, sex, and BMI adjustment, spanning the glycemic markers (fasting glucose, HbA1c, HOMA-IR, and all CGM metrics), the triglyceride-pathway markers (triglycerides, VLDL-C, and the triglyceride-to-HDL ratio), HDL cholesterol, high-sensitivity C-reactive protein, and homocysteine. The low-density-lipoprotein-burden markers (LDL-C, apolipoprotein B, and non-HDL cholesterol) separated the deciles in unadjusted comparisons but carried adjusted standardized coefficients near zero, indicating their separation was largely accounted for by the body-mass term the estimate encodes.

#### 4. Supplementary figures

The supplementary figures are reproduced here with full legends (Figures S1–S5 are referenced in the main text; Figures S5 and S6 document the estimate's development in §1.1). Each is also available as a high-resolution image file.

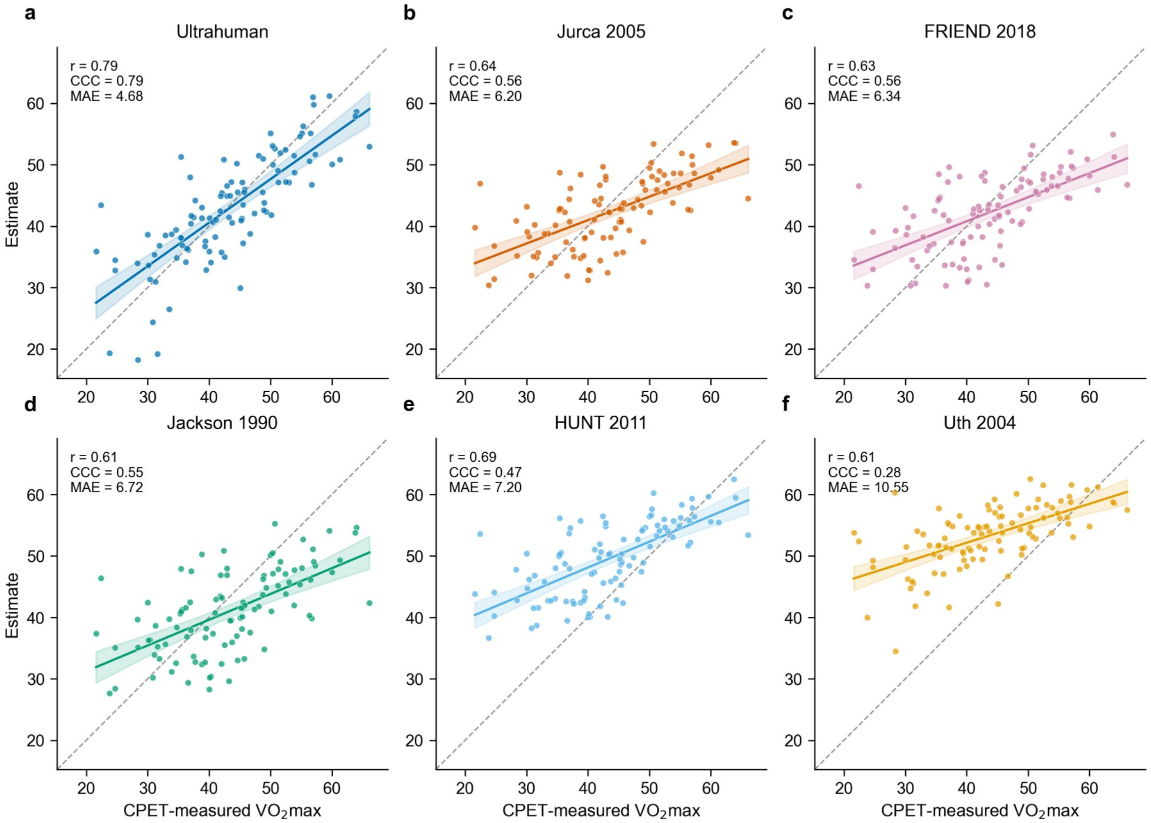


**Figure S1.** Per-formula agreement small-multiples (2×3): agreement scatters against CPET-measured VO2max for the Ultrahuman estimate and the five published equations (Uth, Jurca, Jackson, FRIEND, and HUNT/Nes), each with the line of identity and the fitted regression line (n=101).

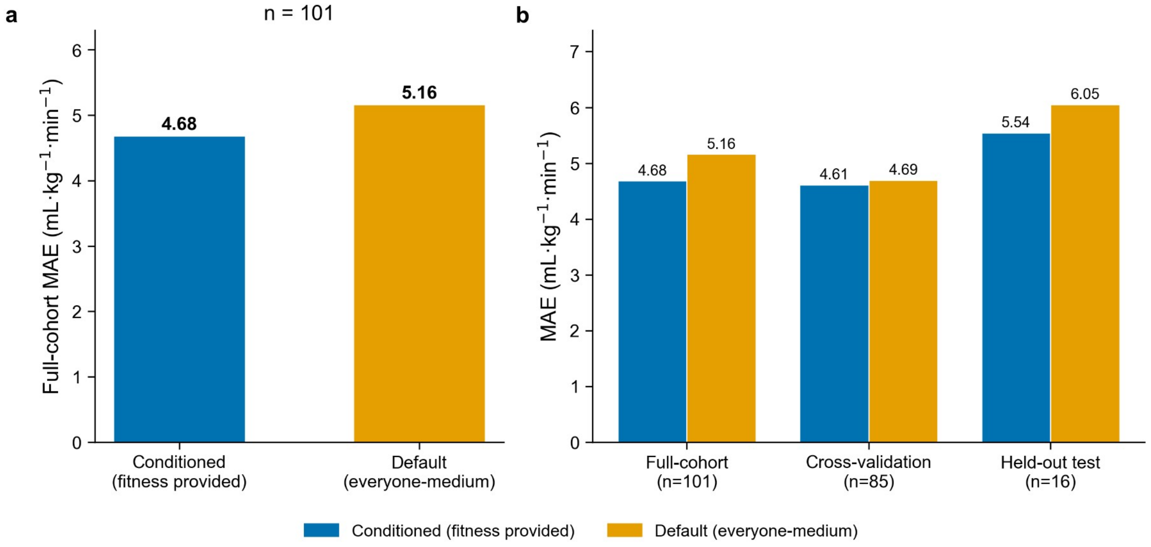


**Figure S2.** Default-configuration (everyone-medium) sensitivity: agreement of the Ultrahuman estimate when the optional self-reported fitness level is absent and every subject is scored on the medium tier (full-cohort MAE 5.16), compared with the conditioned headline result (MAE 4.68), with the cross-validation and held-out-test medium rows.

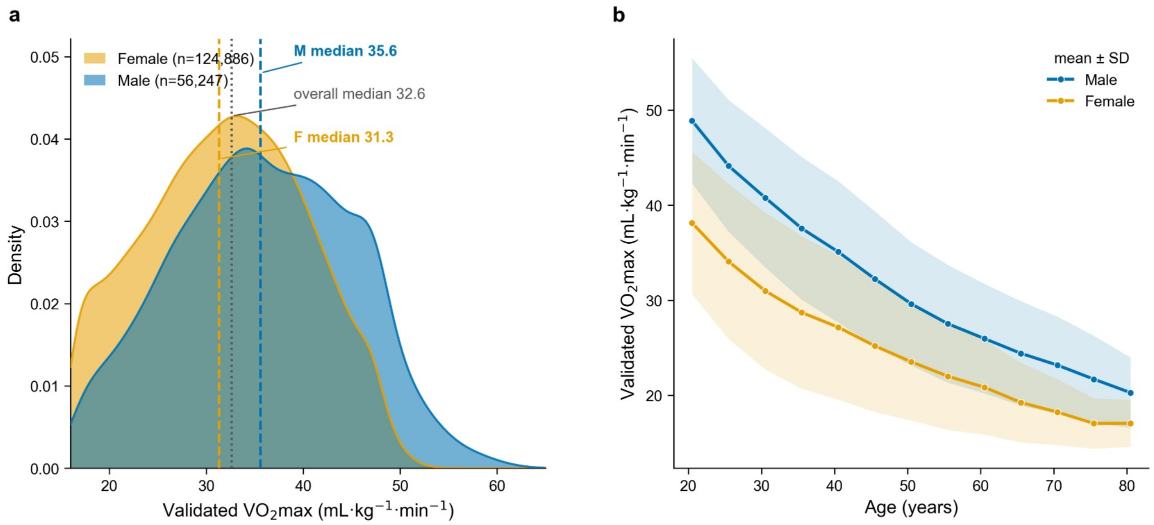


**Figure S3.** Support for the population binning (N=181,133). (a) Smoothed distribution of the validated Ultrahuman VO2max estimate by sex (per-sex Gaussian kernel density; overall median 32.6 mL·kg$^{-1}$·min$^{-1}$, male 35.6, female 31.3), with a low-fitness floor (the ~5.1% of users at the ≤16 mL·kg$^{-1}$·min$^{-1}$ floor) omitted so it does not distort the panel. (b) Validated VO2max declines with age (per-sex binned mean ±1 SD), the relationship that motivates ranking users on the within-sex, age-adjusted residual of the estimate so that the fitness deciles are balanced on age and sex.

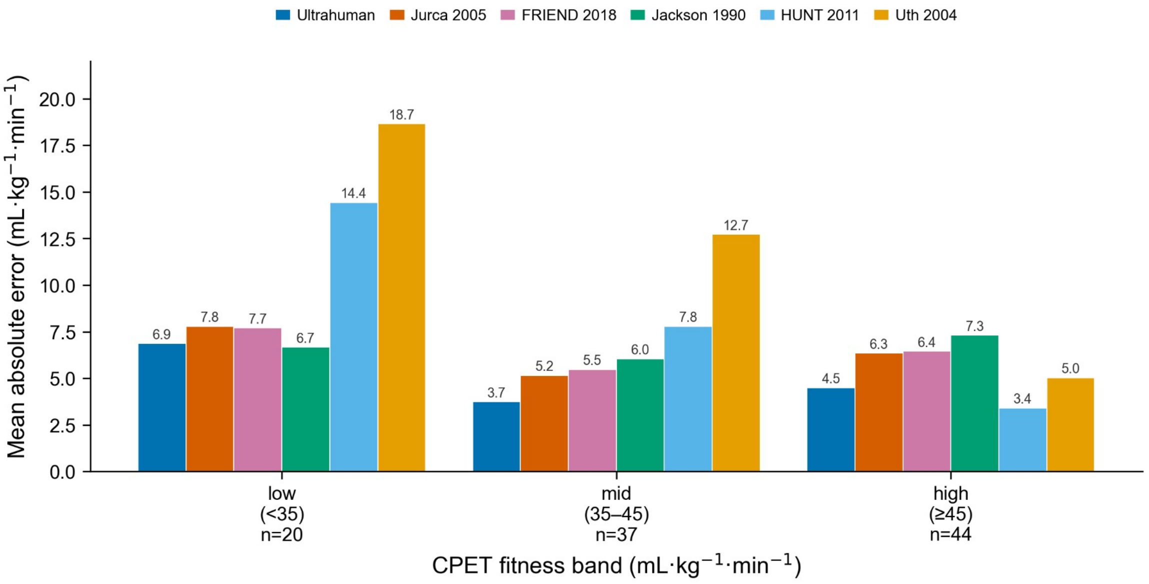


**Figure S4.** Segmented band-level mean absolute error across the low, medium, and high CPET-fitness bands for the Ultrahuman estimate and the five published prediction equations (Uth, Jurca, Jackson, FRIEND, and HUNT/Nes), showing the heteroscedasticity of residuals across the fitness range (n=101).

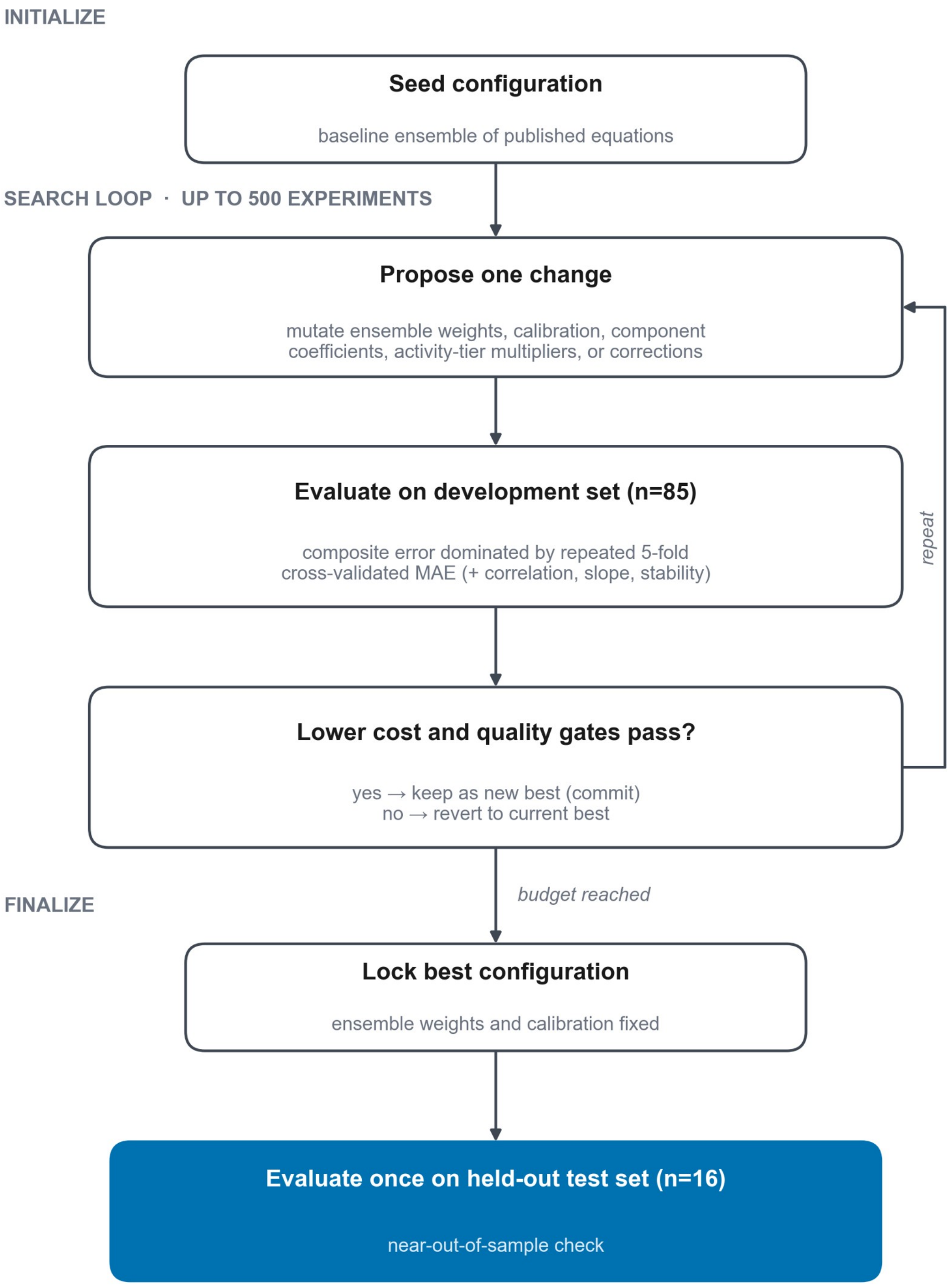


**Figure S5.** Schematic of the autoresearch configuration search used to develop the Ultrahuman Ring AIR VO2max estimate, adapted from Andrej Karpathy's open-source autoresearch loop [5]. After seeding a baseline ensemble configuration, an automated agent repeatedly proposes one change to the configuration (the ensemble weights, the calibration, the per-component coefficients, the activity-tier multipliers, or optional structural corrections), scores the candidate on the 85-subject development set against a composite error criterion dominated by repeated 5-fold cross-validated mean absolute error, and keeps the change only when the cost improves and pre-set quality gates pass, otherwise reverting to the current best; the loop runs to a fixed budget of 500 experiments. The best-scoring configuration is then locked and evaluated once on the 16-subject held-out test set. The diagram shows the process only; the calibrated weights, coefficients, gate thresholds, and offset magnitudes are proprietary (Methods).

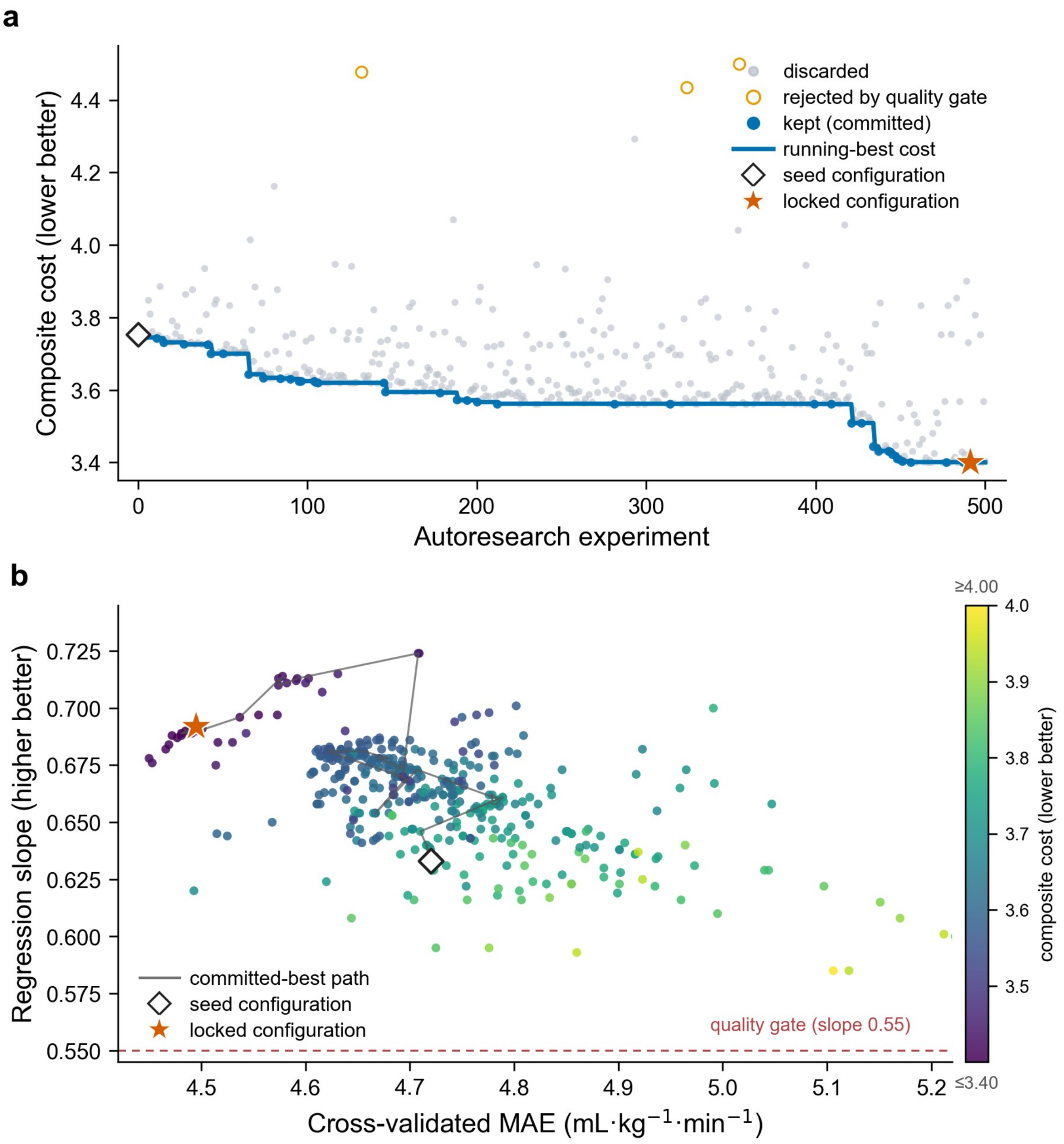


**Figure S6.** How the autoresearch search arrived at the published configuration, from the per-experiment search ledger. (a) Optimization trace: the composite figure of merit (cost) for every scored experiment (discarded, gated, or kept), with the running-best committed configuration descending from the seed (3.7534) to the locked configuration (3.4002), a 9.4% reduction over 500 experiments (41 kept; the final configuration is starred). (b) The same experiments in the plane of cross-validated MAE (x; lower is better) and regression slope (y; 1.0 = no range compression), colored by composite cost, with the seed (open diamond) and the locked configuration (star) marked and the committed-best path traced between them. The search moved toward higher slope and concordance more than toward lower MAE, the agreement axis this validation emphasizes. The composite cost weights several terms, so panel (b) is a projection of the objective and not a Pareto front; the selected configuration is the composite-cost minimum and need not lie on any two-dimensional frontier. Cost is the unitless composite criterion from the search harness (repeated 20-seed nested 5-fold cross-validation on the development set; the 16-subject held-out set excluded). Metrics are the search-time values on the original development corpus; the manuscript's agreement statistics (Table 3) are recomputed on the refreshed cohort, so absolute values differ slightly. Twenty experiments that errored without metrics are omitted, and seven were rejected by a hard quality gate (for example, a regression slope below 0.55). Underlying per-experiment data are provided with the analysis code.

9. Jurca R, Jackson AS, LaMonte MJ, Morrow JR, Blair SN, Wareham NJ, Haskell WL, van Mechelen W, Church TS, Jakicic JM, Laukkanen R. Assessing cardiorespiratory fitness without

performing exercise testing. American Journal of Preventive Medicine 2005;29(3):185-193. PMID:16168867 doi:10.1016/j.amepre.2005.06.004

19. Ye X, Sun M, Yu S, Yang J, Liu Z, Lv H, Wu B, He J, Wang X, Huang L. Smartwatch-based maximum oxygen consumption measurement for predicting acute mountain sickness: diagnostic

accuracy evaluation study. JMIR Mhealth Uhealth 2023;11:e43340. PMID:37410528 doi:10.2196/43340
