## Supplementary material for "Accuracy of a Smart-Ring VO2max Estimate and Five Published Prediction Equations Against Cardiopulmonary Exercise Testing: Development and Validation Study With Population-Scale Analysis": TRIPOD Reporting Checklist

### TRIPOD Reporting Checklist (Multimedia Appendix 2)

**Guideline:** TRIPOD: Transparent Reporting of a multivariable prediction model for Individual Prognosis Or Diagnosis (Collins GS, Reitsma JB, Altman DG, Moons KGM, 2015) [29]

**Applicable model-development phase:** The Ultrahuman Ring AIR VO2max estimate is a multivariable prediction model whose ensemble weights and calibration were selected within this study, on an 85-subject development set drawn at random from the 101-subject cohort, and then evaluated on the 16 held-out subjects. The present study is therefore its **development and validation using a random split-sample (TRIPOD type 2a)**, not an external validation; TRIPOD's Explanation and Elaboration document is explicit that randomly splitting a single data set is a weak form of internal validation rather than external validation. The five published comparator equations, which are applied as published with no fitting to this cohort, are validated here as existing models (**TRIPOD type 4**). Because both development and validation are reported, the development items (10a, 10b, 14a, 15a, 15b) and the validation items (10c, 10e, 12, 13c, 17, 19a) all apply (item 14b, a development item, is in the checklist below but marked not applicable, because no candidate-predictor screen was performed); the departure from item 15a, namely that the fitted ensemble weights and calibrated coefficients are proprietary and are not disclosed, is declared explicitly below and in the manuscript's Limitations. The estimate carries a population-level calibration offset (a policy-tunable population dial, not a per-user fit); re-estimating that offset for a new population (calibration-in-the-large) is identified in the Discussion as the principal generalizability direction rather than something performed within this validation. TRIPOD item numbers and the "Development (D) / Validation (V)" applicability column follow the published checklist.

Locators below name the section headings of the manuscript.

| # | D/V | TRIPOD item | Where addressed in manuscript | Status |
|---|---|---|---|---|
| **Title and Abstract** | | | | |
| 1 | D;V | Identify the study as developing/validating a multivariable prediction model, the target population, and the outcome to be predicted. | **Title**: "Accuracy of a Smart-Ring VO2max Estimate and Five Published Prediction Equations Against Cardiopulmonary Exercise Testing: Development and Validation Study With Population-Scale Analysis." Names development and validation, the target (VO2max / peak VO2), the five comparator equations, and the CPET reference. | Reported |
| 2 | D;V | Provide a structured | **Abstract**: | Reported |

| | | | | |
|---|---|---|---|---|
| | | summary of objectives, study design, setting, participants, sample size, predictors, outcome, statistical analysis, results, and conclusions. | Background / Objective / Methods (N=101 paired ring-CPET cohort, predictors, transparent ensemble structure, 85/16 development/held-out split) / Results (MAE, r, CCC, slope, held-out test set, population-scale construct validity against independent cardiometabolic measures) / Conclusions. | |
| **Introduction** | | | | |
| 3a | D;V | Explain the medical context and rationale for developing/validating the model, including references to existing models. | **Introduction**: mortality rationale for VO2max, CPET as reference, range-compression weakness of wearable estimators, prior-model context (INTERLIVE review; flagship-smartwatch CPET validation). | Reported |
| 3b | D;V | Specify the objectives, including whether the study describes development, validation, or both. | **Introduction** (aims): three aims: quantify agreement, benchmark vs five published equations, assess generalization and population behavior/construct validity. **Statistical analysis** (Methods) states the estimate is a multivariable prediction model, that for the Ultrahuman estimate the study is its development and validation on a random split-sample (TRIPOD type 2a) rather than an external validation, and that the five published equations are validated as existing models (TRIPOD type 4). | Reported |
| **Methods: Source of data** | | | | |
| 4a | D;V | Describe the study design or source of data (e.g., randomized trial, cohort, registry data), separately for development and validation if applicable. | **Study design and participants**: single-site, retrospective observational analysis using de-identified records from adults who underwent a paired CPET-and-ring assessment at the Ultrahuman Performance Lab (Bangalore, India). | Reported |
| 4b | D;V | Specify the key study dates, including start of accrual; end of accrual; and, if applicable, end | **Study design and participants**: CPET assessments were conducted between | Reported |

| | | | | |
|---|---|---|---|---|
| | | of follow-up. | January 11 and June 10, 2026. | |
| **Methods: Participants** | | | | |
| 5a | D;V | Specify key elements of the study setting (e.g., primary care, secondary care, general population), including number and location of centres. | **Study design and participants**: single site, Ultrahuman Performance Lab, Bangalore, India. | Reported |
| 5b | D;V | Describe eligibility criteria for participants. | **Study design and participants**: an age of 18 years or older, a valid CPET peak VO2 measurement, and a concurrent ring-derived night-time resting heart rate (the latter two are the inputs the agreement analysis depends on). | Reported |
| 5c | D;V | Give details of treatments received, if relevant. | No treatment is administered; this is a retrospective observational validation of a passive estimate against a reference test. | Not applicable (observational validation, no intervention). |
| **Methods: Outcome** | | | | |
| 6a | D;V | Clearly define the outcome that is predicted by the prediction model, including how and when assessed. | **Reference test: cardiopulmonary exercise testing**: reference = CPET peak oxygen uptake (VO2peak, $mL \cdot kg^{-1} \cdot min^{-1}$), defined as the highest 30-second rolling average, from a graded treadmill or cycle-ergometer test to volitional exhaustion with breath-by-breath gas exchange (Quark CPET, COSMED, Rome, Italy; OMNIA v2.4.2; gas analysers calibrated daily against a certified reference gas, flow/volume calibrated weekly with a 3-L syringe, ambient temperature/humidity/barometric pressure recorded and applied; treadmill ramp specified; cycle ramp individualized by the Jones-Makrides equation). | Reported |
| 6b | D;V | Report any actions to blind assessment of the outcome to be predicted. | **Statistical analysis**: CPET peak VO2 was determined by the Performance Lab independently of the ring data, and each participant's estimate is computed from that | Reported |

| | | | | |
|---|---|---|---|---|
| | | | participant's ring signals alone, without access to their CPET value, so the index estimate is algorithmically independent of the reference at the per-participant level. | |
| **Methods: Predictors** | | | | |
| 7a | D;V | Clearly define all predictors used in developing/validating the model, including how and when measured. | **The Ultrahuman VO2max estimate** (*inputs*): night-time resting heart rate (photoplethysmography during sleep), demographics (age, sex, weight, height, combined as BMI where used), daily ambulatory activity (step count and a derived mobility tier), and an optional self-reported fitness level with a population-default fallback. | Reported |
| 7b | D;V | Report any actions to blind assessment of predictors for the outcome and other predictors. | Predictors are passively sensor-derived (RHR, steps) or demographic, so no subjective rater assessment applies. **Comparator estimators** and **Multimedia Appendix 1 (§1.2, §1.3)** disclose the input substitutions and imputations carried uniformly across methods. The height plausibility screen (140 to 210 cm) is outcome-blind by construction: the bound was specified a priori from adult biology and applied without reference to the CPET result or to any estimator's error, so it cannot select for participants a given method happens to predict poorly. | Reported (predictors are objective/passive; no rater blinding applicable; the one data-quality screen applied to a predictor is outcome-blind and pre-specified). |
| **Methods: Sample size** | | | | |
| 8 | D;V | Explain how the study size was arrived at. | **Statistical analysis**: no a-priori sample-size calculation was performed; all available paired assessments were included, and achieved precision is reported as bootstrap confidence intervals around each metric. Split sizes (85 development / 16 held-out) are stated in the | Reported |

| | | | | |
|---|---|---|---|---|
| | | | same section; the small-N constraint is noted in **Limitations**. | |
| **Methods: Missing data** | | | | |
| 9 | D;V | Describe how missing data were handled (e.g., complete-case, single imputation, multiple imputation) with details of any imputation method. | Three mechanisms, each specified. (i) *Eligibility*: complete-case on the two inputs the agreement analysis depends on (valid CPET peak VO2 plus a concurrent ring-derived night-time RHR), stated in **Study design and participants**. (ii) *Biological-plausibility screen (missing-data mechanism)*: **Study design and participants** specifies an a-priori adult height bound of 140 to 210 cm, set from the biology rather than from the data and applied to every record without reference to the CPET result or to any estimator's error. One recorded height, 127 cm at a recorded weight of 69 kg (implied BMI 42.8), fell outside it and was set to missing as a data-entry error; the participant was retained on every other input. Height is therefore missing for 2 of the 101 participants (n=99): one never recorded, one set to missing by the screen. The two estimator families then diverge, and the divergence is disclosed in **Multimedia Appendix 1 (§1.2)**: no published equation specifies a missing-height rule, so each comparator was supplied the sex-specific cohort median (173.75 cm, computed over participants with a usable height, so the erroneous value does not enter it), whereas the Ultrahuman estimate applies the missing-height path it ships with (neutral BMI, stature term zeroed) rather than the imputed median. The cohort-level sensitivity | Reported |

| | | | | |
|---|---|---|---|---|
| | | | to that asymmetry is reported in **Multimedia Appendix 1 (§1.2)**: the margin over the best published equation is 1.52, 1.51, and 1.49 mL·kg⁻¹·min⁻¹ under the reported rule, median imputation, and complete-case analysis respectively, and the method ranking is identical in each. A sensitivity analysis retaining the 127 cm value and recomputing every method is reported in **Multimedia Appendix 1 (§1.3)**; no conclusion changes. (iii) *Missing optional predictor*: self-reported fitness is present for 63/101, with the remaining 38 scored on the medium-tier default (**The Ultrahuman VO2max estimate**, **Cohort and peak VO2 distribution**, **Default-configuration result**), and the cost of that fallback is quantified as a bound. | |
| **Methods: Statistical analysis** | | | | |
| 10a | D | Describe how predictors were handled in the analysis. | **The Ultrahuman VO2max estimate**: predictors enter via three published ensemble components (Uth heart-rate ratio, HR-reserve/ACSM, Jackson non-exercise regression) evaluated on a Tanaka age-predicted maximum heart rate deliberately decoupled from same-day activity. | Reported (development-side; the ensemble components are pre-specified published equations, while their weights and calibration were selected within this study). |
| 10b | D | Specify type of model, all model-building procedures (including any predictor selection), and method for internal validation. | **The Ultrahuman VO2max estimate** and **Statistical analysis**: a transparent weighted ensemble of published equations plus a population-level calibration offset and EMA smoothing, with weights/coefficients/offset selected by an automated configuration search (autoresearch loop) minimizing a composite criterion dominated by repeated 5-fold cross-validated | Reported at the disclosed altitude (structure and search procedure fully described; fitted constants proprietary, mirroring Apple/Firstbeat precedent). |

| | | | | |
|---|---|---|---|---|
| | | | MAE on the 85-subject development set (**Use of Artificial Intelligence**; Multimedia Appendix 1, §1.1). Final calibrated constants are proprietary. | |
| 10c | V | For validation, describe how the predictions were calculated. | **Comparator estimators** and **The Ultrahuman VO2max estimate**: how the index estimate and each of the five comparator predictions are computed from the same ring-derived inputs. | Reported |
| 10d | D;V | Specify all measures used to assess model performance and, if relevant, to compare multiple models. | **Statistical analysis**: MAE, mean bias, Pearson r, OLS slope, Lin's CCC, proportion within 5 $mL·kg^{-1}·min^{-1}$, and Bland-Altman limits of agreement; bootstrap 95% CIs (5000 resamples, seed 42). | Reported |
| 10e | V | Describe any model updating (e.g., recalibration) arising from the validation, if done. | **The Ultrahuman VO2max estimate** describes the population-level calibration offset (a policy dial, not a per-user fit); **Out-of-sample generalization** evaluates the locked estimate on the held-out test set, and **Limitations** frames re-estimating that offset for a new population (calibration-in-the-large) as the principal generalizability direction. | Reported |
| **Methods: Risk groups** | | | | |
| 11 | D;V | Provide details on how risk groups were created, if done. | The predicted outcome is continuous VO2max; no risk-group dichotomization of the prediction. Mobility tiers are an input covariate, and the population fitness deciles are a construct-validity grouping, not a predicted risk stratum. | Not applicable (continuous outcome; no risk-group creation for the prediction). |
| **Methods: Development vs validation** | | | | |
| 12 | V | For validation, identify any differences from the development data in setting, eligibility criteria, outcome, and predictors. | **Statistical analysis** and **Out-of-sample generalization**: the development and held-out subsets are random subsets of one cohort, so by construction they do not differ in setting, | Reported (within-cohort validation; separate-population external set named as future work). |

| | | | | |
|---|---|---|---|---|
| | | | eligibility criteria, outcome definition, or predictors. **Limitations**: single-site, Indian (South-Asian) cohort; transport to other populations discussed as re-estimating the calibration level, with a separate-population external set named as future work. | |
| **Results: Participants** | | | | |
| 13a | D;V | Describe the flow of participants through the study, including number of participants with and without the outcome and, if applicable, a summary of follow-up time. A diagram may be helpful. | **Study design and participants** (the analysis cohort comprised 101 evaluable adult participants; eligibility = age 18 years or older, a valid CPET peak VO2, and a concurrent ring-derived night-time RHR) and **Cohort and peak VO2 distribution** (101 analyzed), with the 85 development / 16 held-out split in **Statistical analysis**, **Out-of-sample generalization**, and **Table 3**; **Population-scale behavior** reports the 181,133-user population cohort. | Partially reported: participant counts are given in Methods and Results; a formal participant-flow diagram for the paired cohort is not included. |
| 13b | D;V | Describe the characteristics of the participants (basic demographics, clinical features, available predictors), including the number of participants with missing data for predictors and outcome. | **Cohort and peak VO2 distribution** and **Table 1**: sex (72 male, 29 female), age 32.6 y (SD 7.5), weight 68.9 kg (SD 12.8; n=101), height 170.2 cm (SD 8.8; n=99), BMI 23.7 kg·$m^{-2}$ (SD 3.1; n=99), mobility tier, and CPET peak VO2 overall and by sex. **Missing data for predictors:** height is missing for 2 of 101 (one never recorded, one recorded value of 127 cm set to missing by the 140 to 210 cm adult plausibility screen; Table 1 footnote b, Methods, and **Multimedia Appendix 1, §1.3**), so height and BMI are reported on n=99; the optional self-reported fitness level is missing for 38 of 101 (present for 63/101) and those participants are scored on the medium-tier | Reported |

| | | | | |
|---|---|---|---|---|
| | | | default. The outcome (CPET peak VO2) is complete for all 101 by eligibility. | |
| 13c | V | For validation, show a comparison with the development data of the distribution of important variables (demographics, predictors, outcome). | **Out-of-sample generalization**: the development (n=85) and held-out (n=16) subsets are random subsets of one cohort, and the manuscript reports their outcome distributions side by side (CPET peak VO2 43.1 $mL \cdot kg^{-1} \cdot min^{-1}$, SD 9.5, versus 44.3, SD 11.8; two-sample t test, P=.65) together with their matched fitness-band composition (low 20.0% vs 18.8%, medium 36.5% vs 37.5%, high 43.5% vs 43.8%). | Reported |
| **Results: Model development** | | | | |
| 14a | D | Specify the number of participants and outcome events in each analysis. | **Statistical analysis** and **Out-of-sample generalization**: full-cohort n=101, development/cross-validation n=85, held-out test n=16; default-configuration n's mirrored. | Reported |
| 14b | D | If done, report the unadjusted association between each candidate predictor and outcome. | No candidate-predictor screen was performed. The ensemble components are three pre-specified published equations, and the configuration search selected weights and calibration over those fixed components rather than selecting predictors. The five published single-equation comparators (the published-formula comparison within **Agreement with CPET**) bracket individual-pathway behavior instead. | Not applicable (no candidate-predictor screen; weights and calibration were selected over pre-specified published components). |
| **Results: Model specification** | | | | |
| 15a | D | Present the full prediction model to allow predictions for individuals (i.e., all regression coefficients, and model intercept or baseline survival at a given time point). | **The Ultrahuman VO2max estimate**: the model structure and every ensemble component are disclosed, and **Comparator estimators** publishes all five comparator formulas in full; the final calibrated ensemble weights, | **Declared deviation.** Model structure and all five comparator formulas are published in full, but the fitted ensemble weights, calibrated coefficients, offset magnitude, activity-tier multipliers, and smoothing constant are proprietary and are not |

| | | | | |
|---|---|---|---|---|
| | | | coefficients, offset magnitude, activity-tier multipliers, and smoothing constant are proprietary, as stated in **Data Availability** and **Conflicts of Interest**. | disclosed. Because the study reports model development (TRIPOD type 2a), item 15a applies in full, so this is a departure from the item rather than an inapplicable item. It is stated in **Data Availability**, **Conflicts of Interest**, and **Limitations**, and follows the disclosure boundary set by comparable manufacturer reports. |
| 15b | D | Explain how to use the prediction model. | **The Ultrahuman VO2max estimate** (*inputs*): passive ring signals plus demographics, no maximal or lab test required; an optional self-reported fitness level refines the estimate, otherwise a population default applies. | Reported |
| **Results: Model performance** | | | | |
| 16 | D;V | Report performance measures (with CIs) for the prediction model. | **Agreement with CPET**: MAE 4.68 (95% CI 3.93–5.49), Pearson r 0.792 (0.715 –0.855), Lin CCC 0.787 (0.703–0.847), slope 0.708, 60.4% within 5, Bland-Altman bias −0.40 (−1.61 to +0.80) with 95% LoA [−12.36, +11.56]. The published-formula comparison within **Agreement with CPET** and **Out-of-sample generalization** give the comparators and splits (Tables 2, 3; Figures 3, 4, 5; per-formula scatters in Figure S1). | Reported |
| **Results: Model updating** | | | | |
| 17 | V | If done, report the results from any model updating (i.e., model specification, model performance). | **Out-of-sample generalization** and **Default-configuration result**: held-out test r 0.836 / slope 0.807 (MAE 5.54), everyone-medium default MAE 5.16 (r 0.763, slope 0.616) as a bound; **Population-scale behavior**: population median 32.6 after the calibration offset (Figure 4, Figure 5, Figure S3). | Reported |

| | | | | |
|---|---|---|---|---|
| **Discussion: Limitations** | | | | |
| 18 | D;V | Discuss any limitations of the study (e.g., non-representative sample, few events per predictor, missing data). | **Limitations**: fitness-tier input gap and conditioned headline, slope < 1 range compression, resting-signal correlation ceiling, heteroscedasticity at the extremes, population landing as a calibration choice, small held-out and cross-validation samples, the peak-VO2 (not plateau-confirmed) reference, measurement-timing and age-predicted max-HR caveats, self-reported demographics, and the single-site South-Asian cohort. The missing-data limitation on the anthropometric predictors (height missing for 2 of 101, one of them via the plausibility screen) is stated in **Study design and participants** and **Table 1** (footnote b) and is bounded by the sensitivity analysis in **Multimedia Appendix 1, §1.3**. | Reported |
| **Discussion: Interpretation** | | | | |
| 19a | V | For validation, discuss the results with reference to performance in the development data, and any other validation data. | **Principal Results** and **Out-of-sample generalization**: held-out test versus cross-validation versus full-cohort; r and slope lift with flat MAE read as generalization rather than overfit. | Reported |
| 19b | D;V | Give an overall interpretation of the results, considering objectives, limitations, results from similar studies, and other relevant evidence. | **Principal Results**, **Comparison with Prior Work**, and **Construct Validity at Population Scale**: dissociation of correlation from agreement, ensemble rationale, first ring-vs-CPET and first Indian-cohort framing, comparison to INTERLIVE/Apple/Firstbeat, and independent cardiometabolic gradients. | Reported |
| **Discussion: Implications** | | | | |
| 20 | D;V | Discuss the potential clinical use of the model and implications | **Conclusions**: improved individual ordering within scope, | Reported |

| | | | | |
|---|---|---|---|---|
| | | for future research. | plus three future directions (multi-region CPET set with a transparently re-fitted offset, opt-in fitness capture, sub-maximal effort channel); the Discussion (**Comparison with Prior Work**) ties the estimate to the downstream cardiovascular fitness-age index [32]. | |
| **Other: Supplementary information** | | | | |
| 21 | D;V | Provide information about the availability of supplementary resources (e.g., study protocol, web calculator, datasets). | **Data Availability**: de-identified per-subject table plus analysis/figure scripts on reasonable request; calibrated coefficients proprietary; individual-level raw records withheld for privacy; completed checklists provided as Multimedia Appendices. | Reported |
| **Other: Funding** | | | | |
| 22 | D;V | Give the source of funding and the role of the funders for the present study. | **Funding Statement** and **Conflicts of Interest**: Ultrahuman Healthcare Pvt Ltd, the device manufacturer and employer of all authors, funded the work; the funder's role and the mitigations (five external published benchmarks, a held-out test set withheld from the configuration search, a pre-specified negative control in the population analysis, and separation of data extraction from statistics) are stated, together with the fact that the analysis is retrospective and was not conducted under a registered protocol. | Reported: funder identity, role, and mitigations are stated; per-author equity is not separately itemized, which is a disclosure choice rather than an omission. |

### Notes on partially reported and not-applicable items

These items are addressed transparently below; none is an outstanding gap requiring an editorial fix.

**Partially reported (with reason):**

1. **Item 13a (participant-flow diagram)**: Participant counts are given in **Study design and participants** (101 evaluable participants) and **Cohort and peak VO2 distribution** (101

analyzed), with the 85 development / 16 held-out split in **Out-of-sample generalization** and **Table 3**, and the 181,133-user population cohort in **Population-scale behavior**; a formal participant-flow diagram for the paired cohort is not included.

2. **Item 15a (full model coefficients):** *declared deviation.* The model structure and all five comparator formulas are published in full (**The Ultrahuman VO2max estimate**, **Comparator estimators**), but the final calibrated ensemble weights, coefficients, offset magnitude, activity-tier multipliers, and smoothing constant are proprietary and are not disclosed (**Conflicts of Interest**, **Data Availability**, **Limitations**). Because this study reports model development (TRIPOD type 2a), item 15a applies in full, so this is a departure from the item and not an inapplicable one. It follows the disclosure boundary set by comparable manufacturer reports.

**Not applicable (with reason):**

- **Item 5c (treatments)**: retrospective observational validation of a passive estimate against a reference test; no intervention is administered.
- **Item 11 (risk groups)**: the predicted outcome is continuous VO2max, with no risk-group dichotomization of the prediction.
- **Item 14b (unadjusted per-predictor associations)**: no candidate-predictor screen was performed. The ensemble components are three pre-specified published equations, and the configuration search selected weights and calibration over those fixed components rather than selecting predictors; the five published single-equation comparators bracket individual-pathway behavior.

All remaining TRIPOD items (1, 2, 3a, 3b, 4a, 4b, 5a, 5b, 6a, 6b, 7a, 7b, 8, 9, 10a–10e, 12, 13b, 13c, 14a, 15b, 16, 17, 18, 19a, 19b, 20, 21, 22) are reported in the manuscript.
