## Supplementary material for "Accuracy of a Smart-Ring VO2max Estimate and Five Published Prediction Equations Against Cardiopulmonary Exercise Testing: Development and Validation Study With Population-Scale Analysis": STARD Reporting Checklist

### STARD Reporting Checklist (Multimedia Appendix 3)

**Guideline:** STARD 2015: Standards for Reporting Diagnostic Accuracy studies (Bossuyt PM, Reitsma JB, Bruns DE, et al., 2015) [30]

**Framing for this study.** STARD is applied as the **secondary** reporting guideline (TRIPOD is primary), for a single-site retrospective validation. Here the *index test* is the smart-ring VO2max estimate (the Ultrahuman Ring AIR estimate, with five published prediction equations as comparator index tests), and the *reference standard* is laboratory CPET peak oxygen uptake (VO2peak). Because the target condition (cardiorespiratory fitness, expressed as peak VO2) is a **continuous** quantity, the study reports continuous-agreement metrics (MAE, bias, Pearson r, OLS slope, Lin CCC, Bland-Altman LoA) rather than a 2×2 sensitivity/specificity table; STARD items that presuppose a dichotomous target or a positivity cut-off are annotated "Not applicable (continuous agreement design)".

Locators below are section headings of the manuscript.

| # | STARD item | Where addressed in manuscript | Status |
|---|---|---|---|
| **Title / Abstract / Keywords** | | | |
| 1 | Identification as a study of diagnostic accuracy using at least one measure of accuracy (e.g., sensitivity, specificity, predictive values, AUC). | **Title**, **Keywords** ("validation"; "concordance correlation coefficient"), and **Abstract**. Agreement/accuracy framing is explicit (CCC, slope, MAE). | Reported (accuracy expressed as continuous agreement, not sensitivity/specificity). |
| 2 | Structured summary of study design, methods, results, and conclusions. | **Abstract**: Background/Objective/Methods/Results/Conclusions with index test, reference standard, N, and agreement metrics. | Reported |
| **Introduction** | | | |
| 3 | Scientific and clinical background, including the intended use and clinical role of the index test. | **Introduction**: VO2max mortality association, access gap, wearable estimation; intended use is a passively collected fitness marker, and the range-compression problem motivates the index test. | Reported |
| 4 | Study objectives and hypotheses. | **Introduction** (three aims) and **Abstract** (Objective). | Reported |
| **Methods: Study design** | | | |
| 5 | Whether data collection was planned before the index test and reference standard were performed (prospective study) or after (retrospective study). | **Study design and participants** (opener: "single-site, retrospective observational analysis ... de-identified records from adults who underwent a paired CPET-and-ring assessment at the Ultrahuman Performance Lab"); reaffirmed in **Ethical** | Reported (retrospective). |

| | | | |
|---|---|---|---|
| | | **Considerations** ("retrospective observational research under the ICMR National Ethical Guidelines"). | |
| **Methods: Participants** | | | |
| 6 | Eligibility criteria. | **Study design and participants**: an age of 18 years or older, a valid CPET peak VO2 measurement, and a concurrent ring-derived night-time resting heart rate. | Reported |
| 7 | On what basis potentially eligible participants were identified (e.g., symptoms, results from previous tests, in a registry). | **Study design and participants**: adults who underwent a paired CPET-and-ring assessment at the Ultrahuman Performance Lab (Bangalore, India). | Reported |
| 8 | Where and when potentially eligible participants were identified (setting, location, dates). | **Study design and participants**: single site, Ultrahuman Performance Lab, Bangalore, India; CPET assessments conducted between January 11 and June 10, 2026. | Reported |
| 9 | Whether participants formed a consecutive, random, or convenience series. | **Study design and participants**: a convenience series. Participants attended the laboratory in two series, 24 in an earlier series (tested January 11 to February 27, 2026) and 77 recruited through an open-call VO2max Challenge campaign for the laboratory's assessment service (tested June 1 to 10, 2026), so enrolment was self-selected rather than consecutive. | Reported (convenience series, self-selected through an open call). |
| **Methods: Test methods** | | | |
| 10a | Index test, in sufficient detail to allow replication. | **The Ultrahuman VO2max estimate** and **Comparator estimators**, with **Multimedia Appendix 1 (§1.1, §1.2)**: inputs, ensemble structure (three published component equations), max-HR decoupling, calibration offset, and EMA smoothing fully described, and all five comparator equations fully specified with input approximations. The final calibrated weights and coefficients are declared proprietary. | Reported (structure replicable and all comparator formulas published in full; calibrated constants proprietary as a stated disclosure choice). |
| 10b | Reference standard, in sufficient detail to allow replication. | **Reference test: cardiopulmonary exercise testing**: Quark CPET (COSMED, Rome, Italy) with OMNIA software v2.4.2; gas analysers calibrated daily against certified reference gas, flow and volume calibrated weekly with a 3-L syringe, ambient temperature/humidity/barometric pressure recorded and applied; treadmill ramp (3-min warm-up 3 km/h 0% | Reported (both ramp protocols and the shared metabolic cart are specified; the per-participant modality was recovered for 99 of 101 participants, 78 treadmill and 21 cycle ergometer, reported in Methods and analysed as a moderator in **Limitations**). |

| | | | |
|---|---|---|---|
| | | grade, then 1% grade with speed from 4 km/h increasing 0.5 km/h every 30 s to exhaustion, 5-min cool-down); individualized cycle-ergometer ramp (Jones-Makrides); VO2peak defined as the highest 30-second rolling average. | |
| 11 | Rationale for choosing the reference standard (if alternatives exist). | **Introduction** and **Reference test: cardiopulmonary exercise testing**: CPET is the accepted reference method for peak VO2, with breath-by-breath gas exchange to volitional exhaustion. | Reported |
| 12a | Definition of and rationale for test positivity cut-offs or result categories of the index test, distinguishing pre-specified from exploratory. | Index test yields a continuous VO2max estimate; no diagnostic positivity cut-off is applied. Descriptive agreement bands ("% within 5 $mL\cdot kg^{-1}\cdot min^{-1}$", low-fitness floor ≤16) are pre-specified in **Statistical analysis** and **Population correlate analysis**. | Not applicable (continuous agreement design): descriptive bands disclosed; no diagnostic cut-off. |
| 12b | Definition of and rationale for test positivity cut-offs or result categories of the reference standard, distinguishing pre-specified from exploratory. | Reference standard (CPET peak VO2) is reported as a continuous value (**Reference test: cardiopulmonary exercise testing**); no positivity cut-off. | Not applicable (continuous agreement design). |
| 13a | Whether clinical information and reference standard results were available to the performers/readers of the index test. | **Statistical analysis**: "The estimate for each participant is computed from that participant's ring signals alone, without access to their CPET value." The per-participant index estimate is therefore independent of that participant's reference result; the ensemble weights and calibration were selected on the development set's CPET values, which is why the held-out set provides the out-of-sample read. | Reported |
| 13b | Whether clinical information and index test results were available to the assessors of the reference standard. | **Statistical analysis**: "CPET peak VO2 was determined by the Performance Lab independently of the ring data." | Reported |
| **Methods: Analysis** | | | |
| 14 | Methods for estimating or comparing measures of diagnostic accuracy. | **Statistical analysis**: MAE, bias, Pearson r, OLS slope, Lin CCC, % within 5, Bland-Altman LoA across six estimators (Ultrahuman plus five published), with the rationale for leading with slope and CCC over r; bootstrap 95% CIs. | Reported |
| 15 | How indeterminate index test or reference standard results were handled. | **Reference test: cardiopulmonary exercise testing**: effort was encouraged to volitional exhaustion and assessed against a respiratory exchange ratio approaching or | Reported |

| | | | |
|---|---|---|---|
| | | exceeding 1.10, a near-maximal heart rate, and a plateau where present; not every participant reached RER 1.10 or a plateau, so the highest attained oxygen uptake (VO2peak) was retained as the reference for every participant. Complete-case eligibility (valid CPET plus RHR) is stated in **Study design and participants**; the population arm requires ≥20 valid RHR nights (**Population correlate analysis**). | |
| 16 | How missing data on the index test and reference standard were handled. | **Study design and participants**: height was screened against an a-priori adult plausibility bound of 140 to 210 cm, specified from the biology rather than from the data and applied to every record without reference to the CPET result or to any estimator's error. One recorded height, 127 cm at a recorded weight of 69 kg (implied BMI 42.8), fell outside the bound and was set to missing as a data-entry error; the participant was retained on every other input, so the screen acts as a missing-data mechanism and not as an exclusion. **Cohort and peak VO2 distribution** and **Table 1** (footnote b): height missing for 2 of 101 (n=99; one never recorded, one set to missing by the plausibility screen), BMI therefore computed on n=99; self-reported fitness missing for 38 (scored on the medium tier). **Multimedia Appendix 1 (§1.2)**: no published equation specifies a missing-height rule, so the five comparators were supplied the sex-specific cohort median (173.75 cm, taken over participants with a usable height), whereas the Ultrahuman estimate takes the missing-height path it ships with (neutral BMI, stature term zeroed); this asymmetry is disclosed there, and the cohort-level sensitivity reported in the same section shows the margin over the best published equation (1.52, 1.51, and 1.49 $mL \cdot kg^{-1} \cdot min^{-1}$) and the method ranking are unchanged under either rule. **Multimedia Appendix 1 (§1.3)**: sensitivity analysis retaining the 127 cm value | Reported |

| | | | |
|---|---|---|---|
| | | and recomputing every method, in which no conclusion changes. **Default-configuration result**: medium-tier fallback for missing fitness, quantified. | |
| 17 | Any analyses of variability in diagnostic accuracy, distinguishing pre-specified from exploratory. | **Out-of-sample generalization** (cross-validation vs held-out splits), **Default-configuration result** (with vs without the fitness input), and **Limitations** with **Figure S4** (heteroscedasticity / band-level error across the fitness range). | Reported (pre-specified per Statistical analysis). |
| 18 | Intended sample size and how it was determined. | **Statistical analysis**: "No a-priori sample-size calculation was performed; all available paired assessments were included, and achieved precision is reported as bootstrap confidence intervals." Achieved N=101 and split sizes (85/16) are reported; small N is acknowledged in **Limitations**. | Reported |
| **Results: Participants** | | | |
| 19 | Flow of participants, using a diagram. | **Cohort and peak VO2 distribution** and **Population-scale behavior** give participant counts (101 evaluable paired assessments; 181,133 population), and **Out-of-sample generalization** with **Table 3** give the 85 development / 16 held-out split. A formal STARD flow diagram is not included. | Partially reported: participant counts are given in Results; a formal participant-flow diagram is not included. |
| 20 | Baseline demographic and clinical characteristics of participants. | **Cohort and peak VO2 distribution** and **Table 1**: sex, age, weight, height (170.2 cm, SD 8.8; n=99), BMI (23.7 $kg \cdot m^{-2}$, SD 3.1; n=99), mobility tier, self-reported fitness coverage, and CPET peak VO2 by sex. Table 1 footnote b records the two missing heights and their causes. | Reported |
| 21a | Distribution of severity of disease in those with the target condition. | **Cohort and peak VO2 distribution**: CPET peak VO2 mean 43.3 (SD 9.9), range 21.6 to 66.1 (about threefold), with by-sex means; distribution shown in **Figure 2**. | Reported (severity = full peak-VO2 distribution across low-to-high fitness). |
| 21b | Distribution of alternative diagnoses in those without the target condition. | Continuous target; there is no "without target condition" group or alternative-diagnosis category. | Not applicable (continuous agreement design). |
| 22 | Time interval and any clinical interventions between index test and reference standard. | **Reference test: cardiopulmonary exercise testing**: the ring was worn during the visit and surrounding days so the estimate corresponds to the | Reported |

| | | | |
|---|---|---|---|
| | | same measurement period as CPET; no interventions between tests. **Limitations**: post-test baseline-timing caveat disclosed. | |
| **Results: Test results** | | | |
| 23 | Cross-tabulation of the index test results (or their distribution) by the results of the reference standard. | **Agreement with CPET** with **Figure 3** (estimate-vs-CPET agreement scatter against the line of identity, and Bland-Altman) and **Table 2** (index-vs-reference distribution per method); per-formula scatters in **Figure S1**. A 2×2 cross-tab is not applicable (continuous). | Reported (continuous analog: scatter, Bland-Altman, per-method table). |
| 24 | Estimates of diagnostic accuracy and their precision (e.g., 95% confidence intervals). | **Agreement with CPET**: Ultrahuman MAE 4.68 (95% CI 3.93 to 5.49), r 0.792 (95% CI 0.715 to 0.855), CCC 0.787 (95% CI 0.703 to 0.847), slope 0.708, LoA [−12.36, +11.56]; the five published equations with CIs. **Out-of-sample generalization** and **Tables 2–3**. Bootstrap 95% CIs (5000 resamples, seed 42). | Reported |
| 25 | Any adverse events from performing the index test or the reference standard. | **Reference test: cardiopulmonary exercise testing**: "No adverse events occurred during any test." The index test is passive ring wear. | Reported |
| **Discussion** | | | |
| 26 | Study limitations, including sources of potential bias, statistical uncertainty, and generalizability. | **Limitations**: conditioned headline / fitness-input gap, slope below 1 range compression, resting-signal correlation ceiling, heteroscedasticity, population landing as a calibration choice, small held-out sample, measurement-timing and age-predicted max-HR caveats, the VO2peak (not plateau-confirmed) reference caveat, and single-region South-Asian generalizability. | Reported |
| 27 | Implications for practice, including the intended use and clinical role of the index test. | **Conclusions** and **Comparison with Prior Work**: improved individual-level ordering vs the reference within scope; relation to the downstream wearable fitness-age index; future directions (multi-region CPET set, opt-in fitness input, sub-maximal channel). | Reported |
| **Other information** | | | |
| 28 | Registration number and name of registry. | **Abstract**: "Trial Registration: Not applicable." **Ethical Considerations**: neither analysis is a clinical trial under the New Drugs and Clinical Trials Rules (2019), so no prospective trial registration applies. | Reported (Not applicable, declared with basis). |

| | | | |
|---|---|---|---|
| 29 | Where the full study protocol can be accessed. | **Data Availability**: analysis and figure-generation scripts and a de-identified per-subject table are available from the corresponding author on reasonable request. A separately archived full study protocol is not cited. | Partially reported: scripts and de-identified data are available on reasonable request; no standalone protocol-access point is named. |
| 30 | Sources of funding and other support; role of funders. | **Funding Statement** and **Conflicts of Interest**: Ultrahuman Healthcare Pvt Ltd (device manufacturer and employer of all authors) funded the work; the funder's role in design, testing, analysis, and manuscript preparation is stated, with mitigations (five independently published benchmark equations, a held-out test set withheld from the configuration search, a pre-specified negative control in the population analysis, and separation of extraction/de-identification analysts from the agreement-statistics analysts; the analysis is retrospective and was not conducted under a registered protocol). | Reported (funder role fully reported with mitigations; per-author equity not separately itemized, a disclosure choice). |

### Notes on partially reported and not-applicable items

**Partially reported (transparency notes).**

- **Item 19 (participant flow):** participant counts are given in Results (101 evaluable paired assessments; 181,133 population records), with the 85 development / 16 held-out split in **Out-of-sample generalization** and **Table 3**; a formal participant-flow diagram is not included.
- **Item 29 (protocol access):** the analysis and figure-generation scripts and a de-identified per-subject table are available from the corresponding author on reasonable request; a standalone archived study protocol is not cited.

**Disclosure boundary (Item 10a).** The index-test *structure* (inputs, the three published component equations, max-HR decoupling, calibration offset, and EMA smoothing) and all five comparator formulas are published in full; the final calibrated ensemble weights and coefficients are declared proprietary under the Apple Cardio Fitness and Firstbeat/Garmin disclosure precedent (Methods; Multimedia Appendix 1, §1.1). This is a stated disclosure choice, so the approach can be understood and critiqued without being reproduced commercially.

**Funder disclosure (Item 30).** The funder's role is reported in full (Ultrahuman Healthcare Pvt Ltd, device manufacturer and employer of all authors) together with its mitigations; per-author equity or stock holdings are not separately itemized, which is a disclosure choice rather than an omission of the funder's role.

**Not applicable (continuous-agreement design).** Items **12a** and **12b** carry no index-test or reference-standard positivity cut-off, because the target condition (peak VO2) is a continuous quantity; **Item 21b** has no "without target condition" group or alternative-diagnosis category.

**Not applicable (not a clinical trial). Item 28** (trial registration) is declared Not applicable on the stated basis that the study is retrospective observational research and not a clinical trial under the New Drugs and Clinical Trials Rules (2019).

All remaining STARD items (1–9, 10a, 10b, 11, 13a, 13b, 14–18, 20, 21a, 22–28, 30) are reported in the manuscript. Two items are partially reported (19, 29), and three are not applicable under the continuous-agreement design (12a, 12b, 21b).
